# Phenotype harmonization and analysis for The Populations Underrepresented in Mental illness Association Studies (the PUMAS Project)

**DOI:** 10.1101/2024.10.02.24314732

**Authors:** Ana M Ramirez-Diaz, Ana M Diaz-Zuluaga, Rocky E Stroud, Annabel Vreeker, Mary Bitta, Franjo Ivankovic, Olivia Wootton, Cole A Whiteman, Hayden Mountcastle, Shaili C Jha, Penelope Georgakopoulos, Ishpreet Kaur, Laura Mena, Sandi Asaaf, André Luiz de Souza Rodrigues, Carolina Ziebold, Charles R. J. C Newton, Dan J. Stein, Dickens Akena, Johanna Valencia-Echeverry, Joseph Kyebuzibwa, Juan D Palacio-Ortiz, Justin McMahon, Linnet Ongeri, Lori B Chibnik, Lucas C Quarantini, Lukoye Atwoli, Marcos L Santoro, Mark Baker, Mateus J. A. Diniz, Mauricio Castaño-Ramirez, Melkam Alemayehu, Nayana Holanda, Nohora C Ayola-Serrano, Pedro G Lorencetti, Rehema M Mwema, Roxanne James, Saulo Albuquerque, Shivangi Sharma, Sinéad B Chapman, Sintia I Belangero, Solomon Teferra, Stella Gichuru, Susan K Service, Symon M Kariuki, Thiago H Freitas, Zukiswa Zingela, Ary Gadelha, Carrie E Bearden, Roel A. Ophoff, Benjamin M Neale, Alicia R Martin, Karestan C. Koenen, Carlos N Pato, Carlos Lopez-Jaramillo, Victor Reus, Nelson Freimer, Michele T Pato, Bizu Gelaye, Loes Olde Loohuis

## Abstract

**Background:** The Populations Underrepresented in Mental illness Association Studies (PUMAS) project is attempting to remediate the historical underrepresentation of African and Latin American populations in psychiatric genetics through large-scale genetic association studies of individuals diagnosed with a serious mental illness [SMI, including schizophrenia (SCZ), schizoaffective disorder (SZA) bipolar disorder (BP), and severe major depressive disorder (MDD)] and matched controls. Given growing evidence indicating substantial symptomatic and genetic overlap between these diagnoses, we sought to enable transdiagnostic genetic analyses of PUMAS data by conducting phenotype alignment and harmonization for 89,320 participants (48,165 cases and 41,155 controls) from four cohorts, each of which used different ascertainment and assessment methods: PAISA n=9,105; PUMAS-LATAM n=14,638; NGAP n=42,953 and GPC n=22,624. As we describe here, these efforts have yielded harmonized datasets enabling us to analyze PUMAS genetic variation data at three levels: SMI overall, diagnoses, and individual symptoms.

**Methods:** In aligning item-level phenotypes obtained from 14 different clinical instruments, we incorporated content, branching nature, and time frame for each phenotype; standardized diagnoses; and selected 19 core SMI item-level phenotypes for analyses. The harmonization was evaluated in PUMAS cases using multiple correspondence analysis (MCA), co-occurrence analyses, and item-level endorsement.

**Outcomes:** We mapped >6,895 item-level phenotypes in the aggregated PUMAS data, in which SCZ (44.97%) and severe BP (BP-I, 31.53%) were the most common diagnoses. Twelve of the 19 core item-level phenotypes occurred at frequencies of > 10% across all diagnoses, indicating their potential utility for transdiagnostic genetic analyses. MCA of the 14 phenotypes that were present for all cohorts revealed consistency across cohorts, and placed MDD and SCZ into separate clusters, while other diagnoses showed no significant phenotypic clustering.

**Interpretation:** Our alignment strategy effectively aggregated extensive phenotypic data obtained using diverse assessment tools. The MCA yielded dimensional scores which we will use for genetic analyses along with the item level phenotypes. After successful harmonization, residual phenotypic heterogeneity between cohorts reflects differences in branching structure of diagnostic instruments, recruitment strategies, and symptom interpretation (due to cultural variation).

## Introduction

Genetic investigations of serious mental illness (SMI), including schizophrenia (SCZ), bipolar disorder (BP), and severe or recurrent major depressive disorder (MDD), have identified numerous rare and common variants contributing to risk for these disorders^1–6^. The data sets that have enabled such findings consist of several hundred thousand cases and matched controls, most of whom have European genetic ancestry. The lack of inclusion of participants with other ancestries reflects historical inequities and has limited opportunities for scientific discovery^7–10^. The Populations Underrepresented in Mental illness Association Studies (PUMAS) project was initiated in 2020 to dramatically boost the inclusion in psychiatric genetics of African and Latin American ancestry populations, by creating a dataset that includes more than 120,000 individuals with these ancestries; diagnosed cases of SMI and matched controls paired with genetic variants across the allele frequency spectrum.

The assignment of diagnoses is central to clinical practice, and the consistency of genetic findings for each SMI diagnosis provides a form of validation for these categories. Nevertheless, these diagnoses are by definition heterogeneous; each category consists of a menu of symptoms and other item-level features that may be present in only a subset of individuals within that category, and that may also occur frequently in other categories^11,12^. This heterogeneity is evident in the observation that variation in quantitative SMI-associated phenotypes occurs in trans-diagnostic patterns and in studies showing that a substantial proportion of the genetic variants associated with SMI contribute to risk for more than one diagnosis^13^. Such findings have stimulated efforts to elucidate the biological underpinnings of SMI heterogeneity through genetic analyses at the level of individual items and other trans-diagnostic phenotypes.

Efforts to conduct trans-diagnostic analyses in psychiatric genetics have had to overcome limitations in the data that have been the mainstay of discoveries in the field. These data are mostly contained in separate datasets for SCZ, BP, and MDD, each of which is an amalgamation of many individual studies consisting of cases ascertained and assessed only for that specific diagnosis^11,14^. Individual studies differ from one another in several ways that contribute to heterogeneity in these datasets^15^. Examples include the diagnostic system that a study employs (criteria for diagnoses may differ substantially between such systems^16–18^); the time when a study was conducted (criteria *within* each system for assigning diagnoses evolved over the approximately two decades during which most genetic studies have been carried out); their setting (the symptom profile of cases in an inpatient unit may be very different from that in a primary care clinic); the approach used to elicit clinical features (ranging from detailed clinician-delivered interviews, to self-report inventories, to brief internet-based questionnaires); and even the timeframe assessed (e.g., past 2 weeks, past month, past year, or lifetime). These sources of heterogeneity are particularly challenging for conducting analyses of clinical features at an item level (e.g., ^19^). Efforts to harmonize such phenotypes across studies in existing datasets have been complicated by the lack of available information on key aspects of the design and data types collected in many of the constituent studies.

PUMAS was designed to address the sources of heterogeneity that impede trans-diagnostic analyses, as described above. All PUMAS cohorts focused on ascertaining and assessing SMI cases across diagnostic categories, and obtained information on all of the above sources of heterogeneity. We describe here the approaches that we have used to harmonize diagnoses, symptoms, and other item-level phenotypic features across the entire PUMAS dataset. Specifically, we aligned over 6,895 item-level phenotypes from all instruments used in the PUMAS Project, including clinical interviews, screeners, symptom questionnaires, and targeted instruments (e.g., COVID questionnaires). We created a catalog of harmonized phenotypes, organized by domain, across different instruments and cohorts and identified item-level phenotypes with matching content across instruments and cohorts. From these, we selected a subset of core transdiagnostic SMI phenotypes (n=19) that we intend to use for genetic analyses. We then aggregated data from 89,320 participants across different PUMAS cohorts and evaluated our phenotype harmonization strategy by (i) examining symptom co-occurrence both within and across cohorts and (ii) exploring relationships between these phenotypes at the symptom, diagnosis, and cohort level. Finally, we assessed symptom endorsement rates by cohort and diagnosis.

## Methods

### Participating Cohorts

In this work, we focus on phenotype harmonization across four PUMAS cohorts in which item-level data and diagnoses are derived from clinical interviews. Figure 1 illustrates the geographical distribution of participant recruitment worldwide by PUMAS cohort, and Table 1 presents the recruitment details and general characteristics of each cohort.

**Figure 1.**
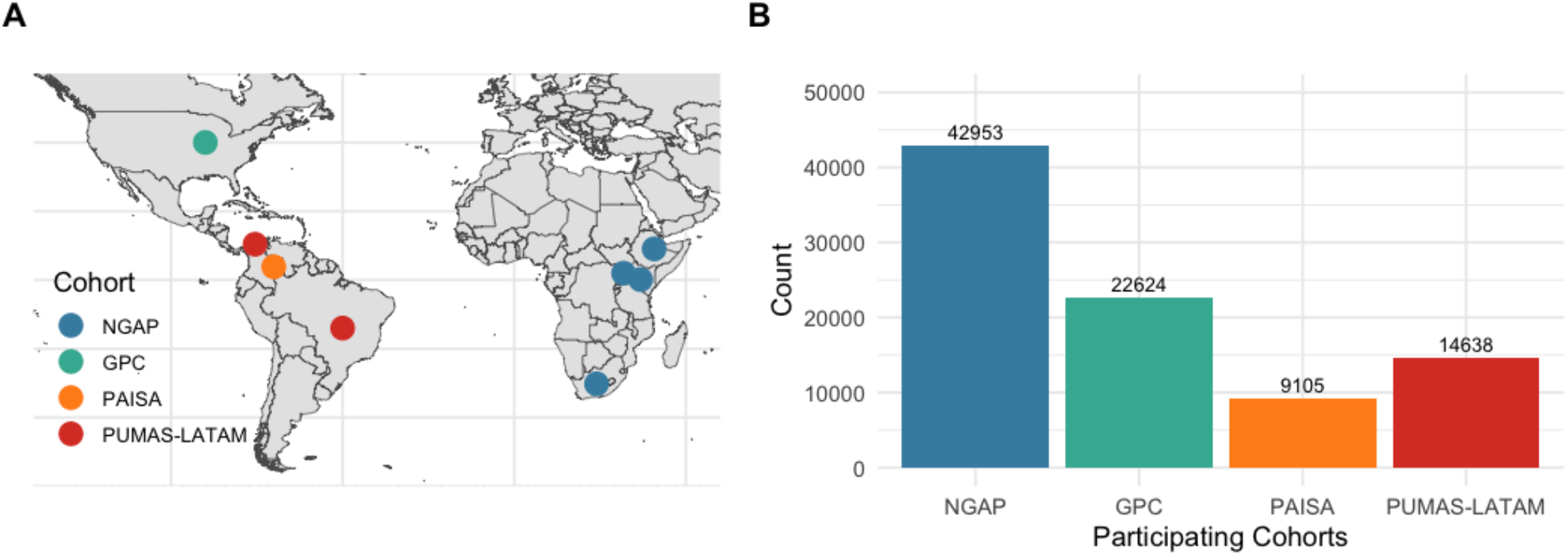
Recruitment countries (A) and sample sizes (B) of PUMAS cohorts, including NGAP, GPC, PAISA, and PUMAS-LATAM. For cohorts with ongoing inclusion (GPC) data freezes up to August 2024 were used (see also Table 1).

**Table 1.** Summary of recruitment details and characteristics of PUMAS cohorts. included in the freeze, including participant country of recruitment, summarized eligibility criteria differences, assessment instruments, screening methods, references to the latest publications for each cohort, data freeze dates, and the total number of participants (N) at the time of data freeze. All cohorts exclusively recruited participants who were 18 years of age or older at the time of enrollment. UBACC = University of California, San Diego Brief Assessment of Capacity to Consent, LEC-5 = Life Events Checklist for Diagnostic and Statistical Manual of Mental Disorders-5, ASSIST = Alcohol, Smoking and Substance Involvement Screening Test, CIDI = Composite International Diagnostic Interview screener, K10 = Kessler Psychological Distress Scale, PSQ = Psychosis Screening Questionnaire, SA-45 = Symptom Assessment-45 Questionnaire, Penn CNB = Penn Computerized Neurobehavioral Battery, Supp. Q = Supplementary Questionnaire.

| Cohort | Country | Eligibility Criteria | Instruments | Screener | Publication (PMID) | Status (Freeze Date) | N |
| --- | --- | --- | --- | --- | --- | --- | --- |
| NGAP | Ethiopia<br>Kenya<br>South Africa<br>Uganda | Cases:<br>Clinical diagnosis of psychosis confirmed by medical record review<br>Capacity to consent to the study as determined by the UBACC | Cases: UBACC, MINI, LEC-5, ASSIST, CIDI, Demographics | - | 30782936<br>35668301<br>36896802 | Completed (03/2023) | 42,953 |
|  |  | Controls:<br>Without Clinical diagnosis of psychosis<br>Capacity to consent to the study as determined by the UBACC | Controls: UBACC, K10, LEC-5, ASSIST, CIDI, Demographics | PSQ |  |  |  |
| GPC | USA | Cases:<br>Clinical diagnosis of Bipolar Disorder or Schizophrenia | Cases: DI-PAD | GPC-S | 23650244 | On going (06/2023) | 22,624 |
|  |  | Controls<br>No lifetime symptoms of psychosis or Mania |  |  |  |  |  |
| PAISA | Colombia | Cases:<br>Must have diagnosis of mood or psychosis spectrum disorder<br>Must meet disease severity criteria, defined by having history of at least one of the following: a hospital admission, treatment for symptoms considered sufficiently severe by a referring psychiatrist to warrant such hospital admission, history of suicide attempt, presence of psychotic symptoms and/or history of electroconvulsive therapy<br>Must have two Paisa last name (to maximize the genetic and cultural homogeneity of the sample) | Cases: NetSCID, Supp. Q, SA-45, Penn CNB, Demographics | EHR and NetSCID screener and overview module | 32353276 | Completed (02/2022) | 9,105 |
|  |  | Controls:<br>Must not have current or lifetime diagnosis of mood or psychotic disorder<br>No current substance use disorder<br>Must have two Paisa last name (to maximize the genetic and cultural homogeneity of the sample) | Controls: NetSCID, Supp. Q, SA-45, Penn CNB, Demographics |  |  |  |  |
| PUMAS LATAM | Colombia<br>Brazil | Cases:<br>Must have diagnosis of mood or psychotic disorder.<br>Must self-report as Mestizo, Black, Amerindian, Hispanic or Latino | Cases: DI-PAD, Treatment Response Questionnaire(Lithium & Antipsyc.), Demographics | GPC-S | - | Completed (07/2024) | 14,638 |
|  |  | Controls:<br>Must not have current or lifetime diagnosis of mood or psychotic disorder in the record<br>Must self-report as Mestizo, Black, Amerindian, Hispanic or Latino | Controls: Demographics |  |  |  |  |
| Total: |  |  |  |  |  |  | 89,320 |

#### NeuroGAP-Psychosis

NeuroGAP-Psychosis (NGAP)^20^ is a case-control study of 42,953 participants recruited across multiple sites within Ethiopia, Kenya, South Africa, and Uganda. Participants included individuals who were at least 18 years old at the time of recruitment and had one of five primary clinical diagnoses (SCZ, schizoaffective disorder (SZA), BP, Psychosis Not Otherwise Specified (NOS) and Mania NOS), and matched controls. Cases were recruited primarily through psychiatric hospitals and community clinics. All cases were interviewed using the Mini-International Neuropsychiatric Interview 7.0.2 Standard (MINI)18. Approximately 7,000 control participants underwent assessment using the MINI as well. Psychiatric diagnoses were established through manual chart review, and item-level phenotypes were extracted from the MINI. NeuroGAP-Psychosis collected additional phenotypes beyond those from this harmonization effort. A full list can be found in the Stevenson et al. protocol paper^20^.

#### Genomic Psychiatry Cohort

The Genomic Psychiatry Cohort (GPC)^21^ is a case-control study of 22,624 participants recruited across multiple states in the United States, including California, Massachusetts, New York, New Jersey, North Carolina, Texas, Ohio, Arizona, Utah, and Maryland. Participants included individuals at least 18 years old at the time of recruitment with a clinical diagnosis of SZA, SCZ, and BP (primarily BP-I) and population controls. Since 2015, the project has focused on patients with self-reported African and Latin Ancestry. Cases were recruited primarily through public university settings. Potential cases and control participants were screened using the GPC screener, a 32-item questionnaire that screens for mania, psychosis, depression, anxiety disorder, alcohol, nicotine, and other substance use history, and medical conditions, including head trauma and seizure history. Controls with lifetime symptoms of psychosis or mania were excluded. All cases were assessed using the Diagnostic Interview for Psychoses and Affective Disorders (DI-PAD)^21^. Psychiatric diagnoses were determined through the Operational Criteria Checklist for Psychotic Illness (OPCRIT) algorithm^22,23^ based on data from the DI-PAD interview. Item-level phenotypes were also extracted from the DI-PAD.

#### Paisa Cohort

The Paisa Project ^24^ is a case-control study of 9,105 participants recruited from the Paisa genetic isolate of Colombia^25–27^. Participants included individuals from the Paisa region who were at least 18 years old at the time of recruitment with a clinical diagnosis of SZA, SCZ, BP, or severe MDD, and matched controls. Cases were recruited from two large psychiatric hospitals in the region. Potential controls were screened using the overview module of the online version of the SCID-5 (NetSCID-5, Spanish Version)^28^. Psychiatric diagnoses were determined through the NetSCID interview. Item-level symptoms were extracted from the i) NetSCID interview, and ii) Supplemental Symptoms Questionnaire (SSQ), an 8-item questionnaire designed to assess transdiagnostic symptoms that the NetSCID’s branching logic did not capture.

#### PUMAS-LATAM Cohort

The PUMAS-LATAM cohort is a case-control study of 14,638 participants recruited from five major cities from different regions in Brazil (São Paulo, Fortaleza, Salvador, Goiânia, Belém), and one coastal city in Colombia (Cartagena). Participants included individuals who were at least 18 years old at the time of recruitment with a clinical diagnosis of SZA, SCZ, and BP, and matched controls. Potential cases and controls were screened with the GPC screener. All cases were assessed using the DI-PAD interview, with psychiatric diagnoses made based on an overall assessment by the recruiting clinician. Item-level phenotypes were also derived from this instrument.

### Phenotype Harmonization Strategy

#### Alignment Procedure

Our phenotype harmonization strategy involved the direct alignment of over 6,895 phenotypes at the level of symptoms and diagnoses drawn from different clinical instruments used across PUMAS cohorts (MINI, NetSCID and DI-PAD), screeners (GPC and NetSCID screening module), and additional targeted instruments (i.e., Alcohol, Smoking and Substance Involvement Screening Test [ASSIST]^29^, Symptom Assessment-45 Questionnaire [SA-45]^30^, Supplementary Symptoms Questionnaire, Substance Abuse Questionnaire, Lithium and Antipsychotic treatment response, COVID questionnaire about impact of the pandemic on mental health, demographic and medical history questionnaires).

First, we collected all clinical instruments and questionnaires used across the different PUMAS cohorts. For each instrument, we had comprehensive documentation of its structure (including branching logic) and administration methodology. Next, building on previous efforts within the Whole Genome Sequencing in Psychiatric Disorders (WGSPD) Consortium^31^, we created a phenotype inventory listing all phenotypes evaluated by each instrument. This inventory captured key attributes of each phenotype, including the assessment timeframe (i.e., current, lifetime, or worst episode), administration method (i.e., self-reported or clinician-administered), and symptom duration (e.g., more than 3 days, 1 week, 2 weeks, more than 2 weeks), and screener, symptom, diagnosis, or specifier item that qualifies other symptom ratings, such as illness duration. We systematically mapped each question from every instrument to its corresponding item-level phenotype, ensuring consistent terminology and conceptual alignment across instruments. Finally, we aligned each individual question by instrument and cohort, while considering branching structure (Supplementary Material 1).

In addition to variability in question phrasing, the response formats used across instruments varied as well: some adopt binary categories (e.g., MINI *Yes/No* options) while others incorporate polytomous response categories (e.g., responses within the DI-PAD included *present, present at least one week, present at least two weeks, present at least one month*). Given the heterogeneity in responses and our primary focus on lifetime symptom endorsement, we dichotomized all responses as “Present” or “Absent.” For instance, NetSCID responses were classified as positive or negative solely at the “threshold” level, and any indication of symptom presence within the DI-PAD was considered as present, regardless of duration. Details on how answer choices were binarized across instruments are listed in Supplementary Material 2.

At the diagnostic level, participating cohorts employed different coding systems, including ICD codes, various DSM classifications and versions, and study-specific designations. Consequently, we harmonized cohort-specific diagnoses into standardized diagnostic categories to ensure consistency: SCZ, SZA, BP, MDD, and Other (Delusional Disorder, Obsessive Compulsive Disorder, Psychotic Disorder Not Otherwise Specified, and Schizophreniform Disorder) (Supplementary Table1). While certain cohorts, such as PAISA and GPC, collected diagnostic information specific to BP type, other cohorts, including NGAP and PUMAS-LATAM, did not differentiate between BP type 1 (BP-I) and other types of bipolar disorder. To identify patients with BP-I among BP patients, in accordance with DSM-5, we defined BP-I as those patients that meet criteria for a lifetime manic episode. All other patients with a diagnosis of BP were classified as “other BP.”

#### Identification of item-level phenotypes

We identified 968 item-level phenotypes that match in content across instruments and cohorts. We selected a subset of 19 item-level phenotypes that are transdiagnostic and core features of SMI. These phenotypes were identified within symptom domains of mania; (grandiosity, decreased need of sleep, flight of ideas, irritability, lifetime manic episode), depression (fatigue, anhedonia, suicide ideation, suicide attempt, suicidality, hypersomnia, sleep disturbances, lifetime depressive episode) and psychosis (delusions, hallucinations, blunted affect, alogia, positive psychotic symptoms and negative psychotic symptoms). Here, positive psychotic symptoms were defined as presence of delusions or hallucinations while negative psychotic symptoms were defined as the presence of blunted or restricted affect, avolition, alogia or abulia. Out of the 19 phenotypes, all but suicide ideation, suicide attempt, blunted affect, alogia, and hypersomnia, were available in all cohorts; 14 phenotypes in total. We created a comprehensive glossary of these phenotypes, detailing the specific instruments’ questions, their branching nature, cohort-specific variable names, the precise phrasing, and the response categories (Supplementary Material 2).

Some phenotypes (suicidality, sleep disturbances, and negative psychotic symptoms) are the result of combining separate items in some instruments. For example, MINI combined questions about suicide attempt and suicide ideation into one item: “Did you repeatedly think about death, have thoughts of killing yourself, or attempt suicide?”, whereas the SCID had separate items for each symptom. Therefore, suicidality includes both suicide attempt and suicide ideation. Similarly, sleep disturbances combine insomnia and hypersomnia, and negative psychotic symptoms include alogia and blunted affect. Details on the specific item-level questions and their branching logic that map to these phenotypes are described in Supplementary Table 2 and Supplementary Material 2.

**Table 2.** Summary Statistics of PUMAS Cohorts including count of participants within each cohort and in total, count and percentage of females, age median and range, count and percentage of controls, count and percentage of females, count and percentage of cases within each diagnostic category, and the percentage of hospitalized participants. Definitions and criteria for each diagnostic category are specified in Supplementary Table 1. NGAP and PUMAS-LATAM did not include participants with MDD (NA), GPC did not include information about Hospitalizations (NA).

| Cohort | NGAP (42,953) | GPC (22,624) | PAISA (9,105) | PUMAS-LATAM (14,638) | Total (89,320) |
| --- | --- | --- | --- | --- | --- |
| Sex (females %) | 18,320 (42.65%) | 10,938 (48.36%) | 5,416 (59.48%) | 7,991 (54.59%) | 42,665 (47.78%) |
| Age: Median [Range] | 35 [18-95] | 43 [18-103] | 44 [18-86] | 39 [18-94] | 38 [18-103] |
| Controls (%) | 21,517 (50.09%) | 11,758 (51.97%) | 1,419 (15.59%) | 6,461 (44.14%) | 41,155 (46.08%) |
| Total Cases (%) | 21,436 (49.91%) | 10,866 (48.03%) | 7,686 (84.41%) | 8,177 (55.86%) | 48,165 (53.92%) |
| BP-I | 6,529 (30.46%) | 2,383 (21.93%) | 2,380 (31.16%) | 3,901 (47.71%) | 15,193 (31.54%) |
| Other BP | 1,068 (4.98%) | 248 (2.28%) | 935 (12.24%) | 496 (6.07%) | 2,747 (5.70%) |
| SCZ | 11,406 (53.21%) | 5,706 (52.51%) | 1,187 (15.44%) | 3,362 (41.12%) | 21,661 (44.97%) |
| SZA | 785 (3.66%) | 1,842 (16.95%) | 320 (4.16%) | 387 (4.73%) | 3,334 (6.92%) |
| Other | 1,648 (7.69%) | 650 (5.98%) | 48 (0.62%) | 31 (0.38%) | 2,377 (4.93%) |
| MDD | NA | 37 (0.34%) | 2,816 (36.64%) | NA | 2,853 (5.92%) |
| History of hospitalization (cases only, %) | 75.63% | NA | 87.31% | 81.33% | 80.06% |

#### Accounting for Branching Logic

We considered the branching logic of each instrument independently, substituting missing values resulting from branching logic with a placeholder of “-9”. Some phenotypes were assessed by multiple questions (e.g., grandiosity had two separate questions on two different modules of the DI-PAD; Supplementary Table 2 and Supplementary Material 2). In these instances, we merged response categories by applying the following hierarchy: 1 (symptom endorsed) > 0 (symptom not endorsed) > -9 (missing due to branching logic) > NA (missing values). This approach allowed us to aggregate symptom ratings across various episodes and instruments, capturing lifetime symptom presence and resulting in binarized responses. We indicate the branching logic of each question in Supplementary Material 1 and 2.

### Data Aggregation

Each site completed individual-level QC, ensuring that all participants met eligibility criteria and had completed the assessment. The aggregated dataset included 89,320 participants from the four participating cohorts. Prior to analyses, we examined data formatting across all variables to ensure adherence to specified format requirements. For the evaluation of phenotype alignment and the phenotypic endorsement analysis, we excluded MDD participants from the GPC cohort due to the limited sample size (n=37). Additionally, participants with “Other” diagnoses—such as delusional disorder, obsessive-compulsive disorder, psychotic disorder not otherwise specified, and schizophreniform disorder—across all cohorts (n=2,377) were not included in the phenotypic analyses to limit heterogeneity. We also excluded controls from item-level analyses due to limited availability of these phenotypes in controls. However, harmonized phenotypes on all participants were generated.

### Evaluation of the Phenotype Alignment

We validated our phenotype alignment through multiple correspondence analysis (MCA)^32^ and co-occurrence analysis of phenotypes in cases. To detect and represent potential phenotypic stratification by cohort and by diagnosis, we used all binary phenotypes to conduct MCA, obtaining dimensional phenotype factors for all participants. For this analysis, we focused solely on the 14 phenotypes available across all cohorts. In addition to detecting underlying structure in phenotype endorsement across cohorts, the variable contributions to the MCA domains also enabled us to confirm the hypothesis that phenotypes within the same domain (psychosis, mania, and depression) load similarly on the MCA dimensions, thus providing additional validation of our approach. In addition to our primary MCA, including all participants and all phenotypes common across cohorts, we evaluated the impact of performing separate MCA by (i) excluding patients with MDD (as these were only included in the Paisa cohort), and (ii) performing MCA on one cohort at a time. To evaluate pairwise phenotypic co-occurrence of phenotypes, we computed the Jaccard Similarity Coefficient for each pair of phenotypes and performed hierarchical clustering on the resulting co-occurrence matrix (using the *hclust* function from the stats package in R).

### Ethics Considerations

This study received approval from the Institutional Review Boards at each participating institution and recruitment site within PUMAS, and all participants signed an informed consent form agreeing to be part of the study.

## Results

### Catalog of Harmonized Phenotypes for Genetic Analysis

We aligned over 6,895 item-level phenotypes, including 14 different clinical instruments, and constructed a catalog of harmonized phenotypes that putatively match in content across different instruments and cohorts. This catalog is organized by domain (i.e., Mood Disorder, Psychotic Disorders, Substance Use Disorder, Anxiety Disorder, Treatment, Demographics) and each item-level phenotype is categorized into screener items (a rating that occurs on a screening section of an instrument, which may result in logical imputation for other items in that domain), symptom items (a symptom of the disorder), and specifier items (a rating that qualifies other symptoms characteristic, e.g., illness duration, seasonal manifestation, age of onset). This catalog aligns not only the terminology of every question from each instrument but also the variable name across studies’ codebooks to facilitate data aggregation and integration. This catalog can be found in Supplementary Material 1.

### Sample characteristics

We successfully aggregated demographic data (age, sex, and country of recruitment), clinical data (diagnosis and history of hospitalization), and 19 phenotypes across 89,320 PUMAS participants (PAISA n=9,105; PUMAS-LATAM n=14,638; NGAP n=42,953 and GPC n=22,624). Our dataset consists of 48,165 cases and 41,155 controls, with 47.78% female and a median age of 38 years. BP-I and SCZ are the most common diagnoses, accounting for 31.54% and 44.97% of the cases, respectively. Table 2 presents demographic characteristics and diagnosis distribution for each cohort and for the total PUMAS cohort.

### Evaluation of Phenotype Alignment

The MCA revealed minimal clustering by cohort along Dimensions 1 and 2 (accounting for 17.63% and 11.04% of the variance, respectively), suggesting a general consistency in phenotypic presentation across cohorts (Figure 2A). An exception is the PAISA cohort, which differs from other cohorts, especially along Dimension 2, and has a notably broader interquartile range along this dimension (1.36) compared to GPC (0.42), NGAP (0.53), and PUMAS-LATAM (0.61). However, these differences are due to the inclusion of MDD patients in this cohort and disappear when MDD patients are excluded from the analysis (Supplementary Figure 1). Excluding patients with MDD reveals a group of NGAP participants that cluster separately (in the bottom right corner of Supplementary Figure 1); further investigation revealed that these are patients that branched out of the MINI’s depression module (Supplementary Figure 2). At the level of diagnosis (Figure 2B), MCA revealed significant symptomatic overlap between patients from different diagnostic categories, emphasizing the non-discrete nature of SMI. At the same time, different diagnostic groups cluster together in expected ways; MDD and SCZ broadly form two separate clusters, while BP-I, other BP, and SZA cover the entire phenotypic spectrum in between, with SZA falling in between BP-I and SCZ. Cohort-specific MCA recapitulate these patterns (Supplementary Figure 3). The component loadings (correlations) of the phenotypes on the first two MCA dimensions group together depressive symptoms (suicidality, sleep disturbances, lifetime depressive episode, anhedonia, and fatigue), manic symptoms (lifetime manic episode, irritability, decreased need of sleep, flight ideas, and grandiosity), and psychotic symptoms (positive psychotic symptoms, hallucinations, delusions, and negative psychotic symptoms).

**Figure 2.**
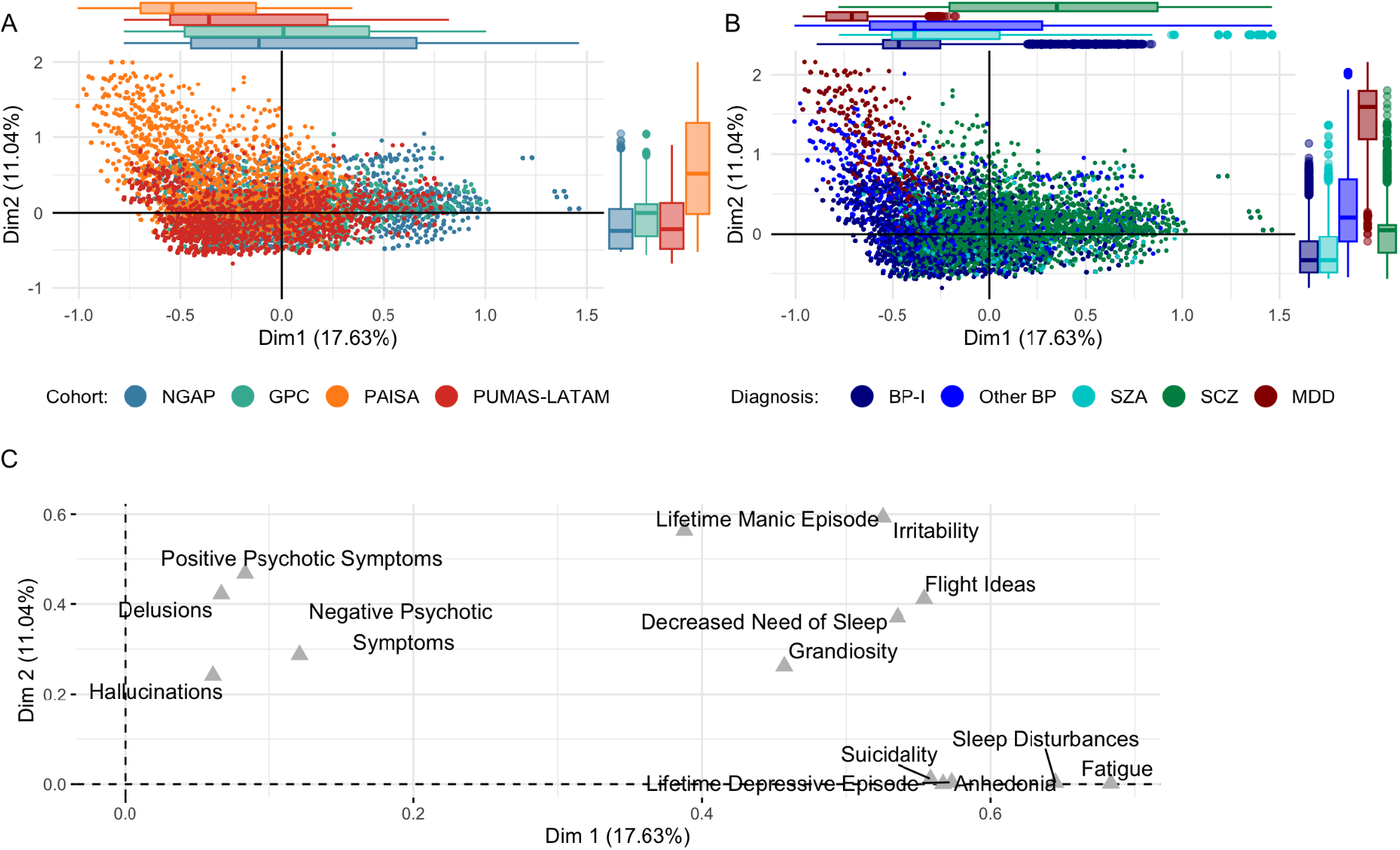
PUMAS-wide Multiple Correspondence Analysis (MCA) (**A & B**) depict the first two dimensions (Dim1 and Dim2) of an MCA performed using item-level phenotypes assessed in all cohorts (positive psychotic symptoms, delusions, hallucinations, negative psychotic symptoms, irritability, flight ideas, grandiosity, lifetime manic episode, lifetime depressive episode, suicidality, anhedonia, sleep disturbances, decreased need of sleep, and fatigue). Individuals are colored by cohort (NGAP, GPC, PAISA, PUMAS-LATAM (**A**) and by Diagnosis (BP-I, Other BP, SZA, SCZ, and MDD) (**B**). The percentage of variation (inertia) explained by each dimension is indicated in parentheses. The marginal box plots summarize the distribution of the MCA scores along Dim1 and Dim2 for each cohort or diagnosis group. The central line in each box represents the median of the data, while the hinges of the box indicate the first and third quartiles. (**C**) Visualizes the variable correlations to the MCA dimensions.

Similarly, the co-occurrence clustering revealed distinct co-occurrence clusters of depressive, mania, and positive psychotic symptom domains (Figure 3). A fourth symptom cluster is the phenotype of negative psychotic symptoms, a core symptom domain of psychotic disorders like SCZ, which formed a cluster on its own. We observed low co-occurrence between negative psychotic symptoms and most other symptoms, with a maximum co-occurrence with positive psychotic symptoms (Jaccard 0.51), and relatively high co-occurrence between depressive and mania symptoms (Figure 3). Grandiosity, while broadly clustering with mania phenotypes (lifetime manic episodes, flight of ideas, decreased need for sleep), displayed markedly lower co-occurrence with other mania symptoms (Jaccard 0.65-0.67) than the remaining mania symptoms with each other (Jaccard 0.72-0.85). We observe a similar pattern with suicidality and depressive symptoms (Jaccard 0.63-0.64 versus 0.80-0.86). Further co-occurrence analyses performed for each cohort separately broadly recapitulate these observations, with tight clusters for psychotic and depressive symptoms and a lack of clustering between negative symptoms and any other phenotypes. Notable variations between the cohorts are observed for the phenotypes of suicidality, irritability, and grandiosity, which, depending on the cohort, cluster together with manic, depressive, or psychotic symptoms (Supplementary Figure 4).

**Figure 3.**
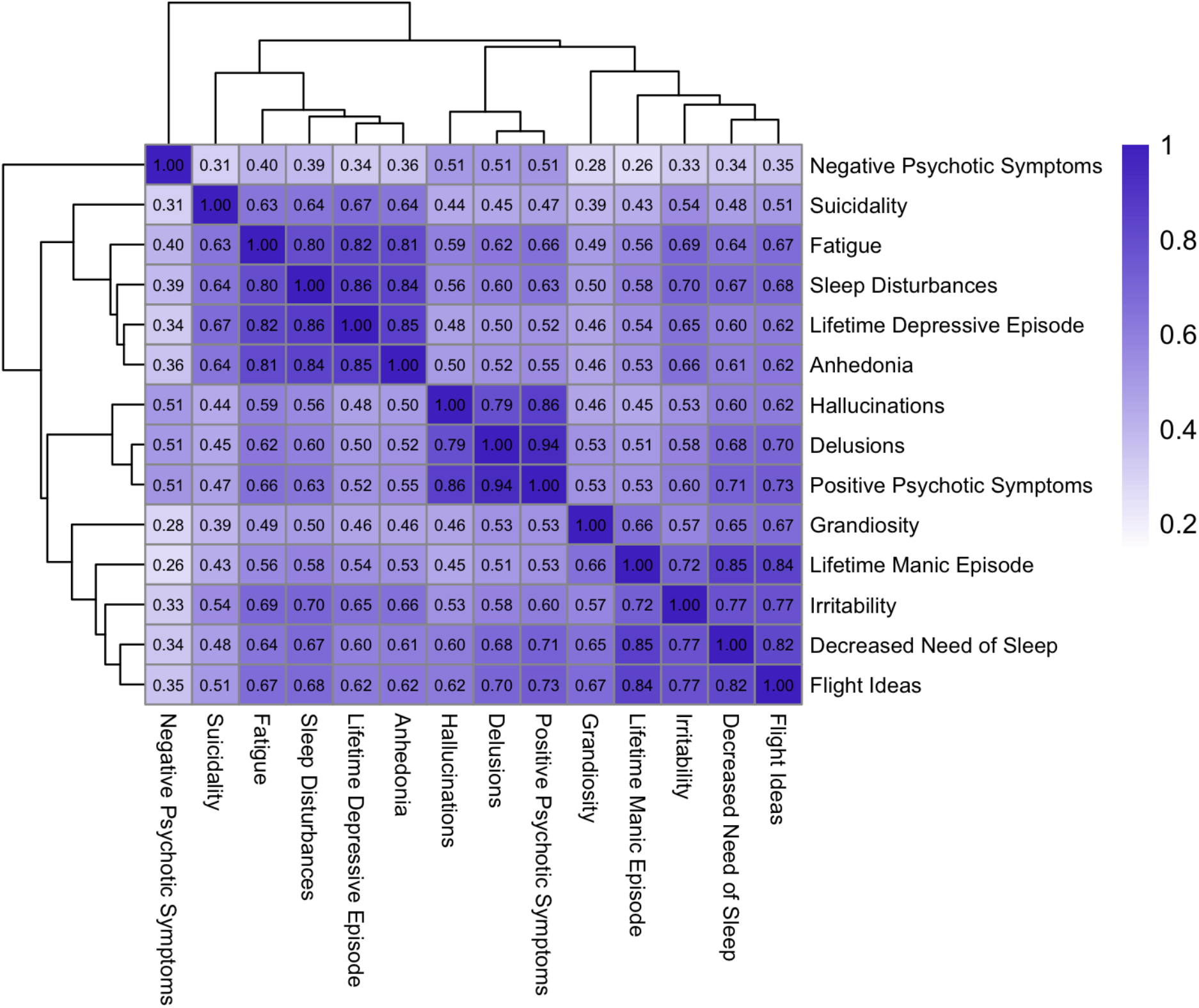
Jaccard similarity matrix of phenotypes including all PUMAS cohorts. Each cell in the matrix corresponds to the Jaccard’s Similarity of two phenotypes, calculated as the proportion of instances where both phenotypes are endorsed relative to the total number of instances where at least one of the phenotypes is endorsed. The gradient scale from light to dark reflects the range of Jaccard Similarity Scores, from 0 (no similarity) to 1 (perfect similarity).

Taken together, these MCA and co-occurrence analyses provide validation that our phenotype harmonization was successful while highlighting the heterogeneous, dimensional, and overlapping nature of SMI.

### Phenotype Endorsement

The item-level endorsement rates indicate that most (12/19) phenotypes were transdiagnostic, with over 10% endorsement across the diagnostic groups of BP-I, Other BP, SZ, MDD, and SZA (Supplementary Table 3). Phenotypes that do not meet this threshold - manic episodes, grandiosity, flight of ideas, delusions, blunted affect, and negative psychotic symptoms – were all uncommon in severe MDD (<10%). Depression-related phenotypes were highly prevalent across diagnoses. For instance, lifetime depressive episode and anhedonia were frequent, with over 73% endorsement across all diagnostic categories, except in SCZ, where the rates were 44.8% and 49.6%, respectively. Additionally, suicidality, suicidal ideation, fatigue, and sleep disturbances were reported by more than 50% of cases across all diagnoses, except for SCZ. The lowest endorsement rate for suicide attempt was in SCZ (23.6%), while MDD had the highest rate (50.1%). Mania-related symptoms, such as irritability, decreased need for sleep, and grandiosity, were predominantly observed in BP-I patients, rates decreasing for Other BP, SZA, SCZ, and least frequently in MDD. Psychotic symptoms, including delusions, hallucinations, and positive psychotic symptoms, were frequently endorsed across all diagnoses, with more than 50% endorsement, except for MDD, where these rates were significantly lower (9.5%, 16.8%, and 19.9%, respectively). Negative symptoms such as blunted affect, alogia, and general negative psychotic symptoms were primarily observed in SCZ patients (62.8%, 48.5%, and 65.1%, respectively). Notably, hypersomnia was one of the lowest endorsed phenotypes, with highest frequency in BP-I patients (39.1%) and lowest in SCZ patients (23.7%).

Overall, item-level phenotypic profiles of diagnostic groups were consistent across cohorts, with some notable variations (Figure 4). Patients with SCZ diagnoses in NGAP endorsed lifetime manic episodes at a higher frequency (45.5%) than other cohorts (GPC 9.4%, PAISA 12.4%, PUMAS-LATAM 16.0%). In comparison to other cohorts, participants from GPC reported higher frequencies of fatigue in BP-I (92.7%), SZA (93.7%), and SCZ patients (72.4%), as well as higher rates of suicidality (BP-I 68.5%, SZA 75.8%, SCZ 40.5%) and suicide attempts (BP-I 48.4%, SZA 56.6%, SCZ 36.1%). In contrast, BP-I and Other BP patients from the PAISA cohort reported lower endorsement of psychotic symptoms compared to patients in this group from other cohorts, while NGAP patients reported lower rates of manic symptoms. Notably, the GPC and PUMAS-LATAM cohorts, both employing the DI-PAD interview, demonstrated the most similar symptom endorsement patterns. An overview of the proportion of patients that branched out of each phenotype, by cohort, is provided in Supplementary Figure 5, highlighting that branching is not only common for depressive symptoms in NGAP, as described above, but also for blunted affect and negative psychotic symptoms in PAISA.

**Figure 4.**
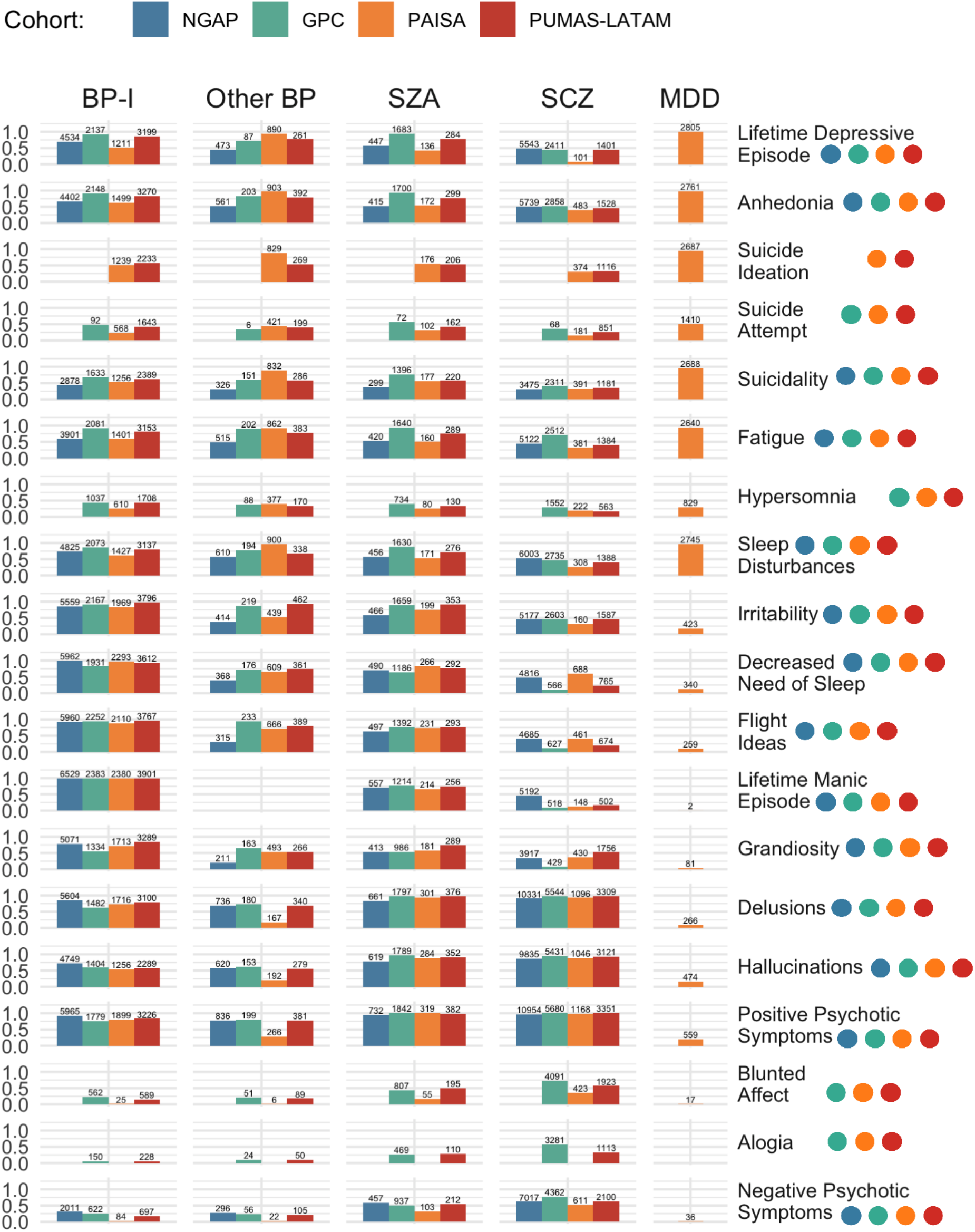
Phenotype endorsement by cohort and diagnosis. Colored dots represent phenotypic data availability per cohort.

## Discussion

We aligned symptoms and diagnoses across several assessment instruments in four cohorts, establishing a framework for trans-diagnostic harmonization of item-level phenotypes in large-scale psychiatric genetics research. In this process, we mapped over 6,895 item-level phenotypes and aggregated data from more than 89,000 individuals of Latin American and African ancestry, including >48,000 with SMI diagnoses. We delineated and analyzed 19 harmonized items that are key components of SMI.

### Variations in frequencies of SMI diagnoses between cohorts

Although none of the PUMAS cohorts could be considered an epidemiological sample, they all represent efforts to ascertain SMI cases in an unbiased and comprehensive way from the population of individuals seeking care for psychotic and bipolar spectrum disorders in their respective locations. From this standpoint, the substantial difference between sites in the proportion of participants with each of these diagnoses is noteworthy. Specifically, SCZ was more frequent than BP-I in NGAP and GPC, while the reverse pattern was observed in PUMAS-LATAM and PAISA. Variations in recruitment settings and protocols, along with cultural and geographic variations in diagnostic practices likely contribute to these patterns. It is also possible, however, that these observations may reflect differences between the recruitment sites in factors contributing to the expression of SMI, i.e., either environmental exposures or genetic variations. Further evaluation of clinical profiles of patients as well as genetic analyses will contribute to our understanding of these differences^33,34^.

### Phenotype harmonization identifies diagnosis-specific and trans-diagnostic patterns in the endorsement of item-level clinical features

Our procedures enabled us to examine these patterns in a unified way across all four PUMAS cohorts. As expected, individual clinical features are most frequently observed in participants with diagnoses typified by these features^18^. However, most features are also observed at substantial frequencies across categories, presenting the opportunity to use them as phenotypes in PUMAS genetic analyses. For example, nearly half of the participants with diagnoses of SCZ endorsed anhedonia, typically considered a manifestation of severe MDD. It is a topic of debate within the field whether such anhedonic symptoms are truly trans-diagnostic or are representations of different phenomena described in similar terms by participants in clinical interviews^35^. Genetic analyses could shed light on this debate, indicating, for example, whether the genetic risk profile of anhedonia in MDD patients is like that of anhedonia among SCZ patients. Additionally, aggregating harmonized trans-diagnostic item-level features across the entire PUMAS dataset will enable common and rare variant discovery by minimizing phenotypic and genetic heterogeneity. Although the assessment methods used in PUMAS primarily involve recording individual symptoms as categorical features, the MCA provides a potential means to incorporate dimensional aspects of SMI in genetic analyses, as called for in the NIMH Research Domain Criteria (RdoC) framework^36^. The first two MCA dimensions show that MDD and SCZ generally form distinct clusters; BP and SZA are distributed across a spectrum between MDD and SCZ. We propose that these MCA dimensions provide a starting point for quantitative genetic analyses of SMI within PUMAS.

### Limitations of phenotype harmonization and future directions

Phenotype harmonization strategies cannot eliminate all sources of heterogeneity. Notably, differences in phenotyping instruments, particularly different branching structures, result in different endorsement rates of various phenotypes that may not underlie clinical differences. For example, among case participants, branching in the depressive module of the MINI, used only in NGAP, likely contributed to a lower endorsement rate of depression items in this cohort compared to other cohorts. Additionally, the four PUMAS cohorts analyzed here are drawn from populations that are culturally quite different from each other. Harmonization strategies cannot fully capture differences in clinical features that are due to site-specific diagnostic practices and cultural variations in symptom interpretation and reporting among patients. It may be possible to shed light on the factors contributing to item-level heterogeneity between cohorts, for example through GWAS analyses that evaluate variations in symptom endorsement across diagnoses and across populations.

Here, we used MCA to summarize the phenotypic variance in our data across cohorts and diagnoses. We aim, in future studies, to extend the psychometric evaluation of our phenotype harmonization approach. For example, identifying latent factors and conducting formal investigations of measurement equivalence (invariance) will help clarify how cohort heterogeneity affects the comparability of these latent factors. Cross-diagnostic genetic analysis of these phenotypes and their dimensional derivatives as part of PUMAS will reveal possible genetic etiology within and across diagnoses and ancestral populations.

## Conclusion

This study provides a framework in psychiatric genetics research for phenotype harmonization of item-level phenotypes, highlighting shared and distinct symptom profiles across different major psychiatric disorders and populations that have been historically underrepresented in psychiatric (genetic) research. By successfully harmonizing psychiatric diagnoses and phenotypes across instruments, cohorts and continents, we identified and effectively aggregated data of 19 transdiagnostic SMI phenotypes for future genetic analysis. The PUMAS Project will enable the dissection of the genetic architecture of specific phenotypes, and their contribution to SMI.

## Supporting information

Supplementary Material 1. PUMAS Catalog of Harmonized Phenotypes for Genetic Analysis

Supplementary Material 2. Glossary of 19 Harmonized Phenotypes

## Data Availability

All data produced in the present study are available upon reasonable request to the authors.

## Acknowledgements

This work is supported by a Collaborative U01 grant from the National Institute of Mental Health (NIMH): Powering Genetic Discovery for Severe Mental Illness in Latin American and African Ancestries awarded to: The Broad Institute of MIT and Harvard (U01MH125047); Harvard T.H. Chan School of Public Health (U01MH125045); Universidad de Antioquia and University of California Los Angeles (U01MH1250452); and Rutgers University (U01MH125049). Participant recruitment and analysis of the PAISA Cohort study was supported through NIMH R01MH113078. Participant recruitment and analysis in the NeuroGAP-Psychosis study was provided by The Stanley Family Foundation as well as R01MH120642.

## Supplementary Material

Supplementary Tables and Figures

Supplementary Material 1. PUMAS Catalog of Harmonized Phenotypes for Genetic Analysis

Supplementary Material 2. Glossary of 19 Harmonized Phenotypes

