## Supplementary Tables and Figures for "Phenotype harmonization and analysis for The Populations Underrepresented in Mental illness Association Studies (the PUMAS Project)"

### Index:

**Supplementary Table 1**. Harmonization of Diagnoses Across Cohorts
**Supplementary Table 2**. Harmonization of Phenotypes Across Cohorts
**Supplementary Table 3.** Phenotype Endorsement proportion by Diagnosis Group

**Supplementary Figure 1.** PUMAS-wide Multiple Correspondence Analysis (MCA) excluding MDD patients from Paisa Cohort.
**Supplementary Figure 2.** PUMAS-wide MCA highlighting NGAP participants cluster that branched out of depression module.
**Supplementary Figure 3.** Cohort-specific Multiple Correspondence Analysis (MCA)
**Supplementary Figure 4.** Jaccard Similarity Matrix of Phenotypic Variables for each PUMAS Cohort (excluding MDD patients from Paisa Cohort)
**Supplementary Figure 5.** Phenotype Endorsement by Cohort including branching

| | GPC | NGAP | PAISA | PUMAS-LATAM |
|---|---|---|---|---|
| **Bipolar Disorder*** | Bipolar I disorder<br>Bipolar I with psychosis<br>Bipolar II disorder<br>Bipolar II with psychosis<br>Manic episode (Bipolar I)<br>Manic episode with psychosis (Bipolar I)<br>Hypomanic Episode/s | Bipolar<br>Mania NOS | 296.40-.46: Bipolar I, current manic/hypomanic episode<br>296.50-.56: Bipolar I, current depressive 296.7: Bipolar I, current unspecified<br>296.80: Unspecified Bipolar Disorder<br>296.89: Bipolar II Disorder | F31.0: Bipolar Disorder- Hypomanic Episode<br>F31.1-F31.2: Bipolar Disorder current manic<br>F31.3-F31.5: Bipolar Disorder current manic<br>F31.6: Bipolar Affective Disorder current mixed F31.6: Bipolar Disorder in remission<br>F30.1-F30.2: Manic Episode |
| **Schizoaffective Disorder** | Schizoaffective disorder -- bipolar type<br>Schizoaffective disorder -- depressive type | Schizoaffective | 295.7: Schizoaffective disorder | F25: Schizoaffective disorder |
| **Schizophrenia** | Schizophrenia<br>Schizophrenia -- Hebephrenic,<br>Schizophrenia -- Paranoid<br>Schizophrenia -- Undifferentiated | Schizophrenia | 295.9: Schizophrenia | F20: Schizophrenia |
| **Major Depressive Disorder** | Major depressive disorder (MDD)<br>Major depressive disorder -- moderate<br>Major depressive disorder – severe<br>MDD with psychosis | | 296.21-296.26: Major depressive affective disorder, single episode<br>296.30-296.36: Major depressive affective disorder, recurrent episode | |
| **Other** | Delusional disorder<br>Obsessive Compulsive Disorder<br>Psychosis NOS<br>Schizophreniform<br>Unaffected Family Members | Psychotic Disorder NOS | 297.1, 311, 625, 298.8-9<br>300.00-09, .22, .23, .29, .30, .90<br>304.00, .10, .30, .40, .60<br>305.00, .20, .70: 307.1, .51, .59<br>309.28, .81, .89, .90* | F22: Delusional Disorder |
| **Controls** | Control, Control: depression. | No Diagnosis | No Diagnosis | No Diagnosis |

**Supplementary Table 1: Harmonization of Diagnoses Across Cohorts** lists the cohort-specific diagnosis that have been mapped into six harmonized Diagnostic Groups: Bipolar Disorder, Schizoaffective Disorder, Schizophrenia, Major Depressive Disorder, and Other, with a separate category for Controls. *Bipolar Disorder Type I (BP-I) definition: To identify patients with BP-I among BP patients, in accordance with DSM-5, we defined BP-I as those patients that meet criteria for a lifetime manic episode. All other patients with a diagnosis of BP were classified as “other BP”.

**Other category in PAISA Cohort includes: Obsessive-compulsive disorder, Unspecified anxiety disorder, Alcohol use disorder, past - Moderate. Without remission. Obsessive-compulsive disorder, Anorexia Nerviosa, Adjustment Disorder, GAD, Disorder induced by substances, Panic disorder, Agoraphobia, Specific Phobia, Bulimia nerviosa, Binge-eating disorder, PTSD, TDAH.

| | GPC | NGAP | PAISA | PUMAS-LATAM |
|---|---|---|---|---|
| **Positive Psychotic Symptoms** | Presence of any delusions/ hallucinations | (Psychosis) Does the participant meet criteria for a Lifetime (or Current) Psychotic Disorder? | Presence of any delusions/ hallucinations | Presence of any delusions/ hallucinations |
| **Lifetime Depressive Episode** | Ask the respondent to estimate the number of depressive episodes they've had. | Does the participant meet criteria for a Past Depressive Episode? | DSM-5 Criteria for Depressive Episode met | Ask the respondent to estimate the number of depressive episodes they've had. |
| **Lifetime Manic Episode** | Ask the respondent to estimate the number of manic episodes they've had. | Does the participant meet criteria for a Past Manic Episode? | DSM-5 Criteria for Manic Episode met | Ask the respondent to estimate the number of manic episodes they've had. |
| **Grandiosity** | Grandiose delusions: Have you ever felt that you had any special power, talents, or abilities -- much more than other people? Have you ever felt that you had a special purpose, mission, or identity? Or that you were rich or famous, or related to prominent people? Or, maybe, that you had been chosen by God for a special mission? Do you believe that this really is true? How long have you felt like this?<br><br>Increased self-esteem: During that time, did you have grand beliefs or ideas that you later found out were not true? Like believing that you had special powers or abilities others did not have? Or that you could accomplish much more at work or in your daily activities, as if you had super powers or talents? How long did you feel like that? | Feel that you could do things others couldn't do, or that you were an especially important person? | **SCID:** During the time of mood disturbance did you feel more confident than usual? Did you feel much smarter or better than everyone else? Did you feel like you had special powers or abilities?<br><br>**SCID:** Did you ever think that you were especially important in some way, or that you had special powers or knowledge?<br><br>**SCID:** Have you ever thought that in some way you were especially important or that you had special powers or knowledge?<br><br>**Supp. Q:** Did you have a period of at least 1 week where you have felt, in an unusual way, more confident in your abilities and capabilities and more competent and talented than other people. your neighborhood? | Grandiose delusions: Have you ever felt that you had any special power, talents, or abilities -- much more than other people? Have you ever felt that you had a special purpose, mission, or identity? Or that you were rich or famous, or related to prominent people? Or, maybe, that you had been chosen by God for a special mission? Do you believe that this really is true? How long have you felt like this?<br><br>Increased self-esteem: Have you ever/ During that time, did you have grand beliefs or ideas that you later found out were not true? Like believing that you had special powers or abilities others did not have? Or that you could accomplish much more at work or in your daily activities, as if you had super powers or talents? How long did you feel like that? |
| **Delusions** | Individual questions asking for the presence of each of following type of delusions:<br>Persecutory/ Jealous<br>Mind reading Thought insertion/ broadcasting/ withdrawal<br>Echo<br>Passivity<br>Influence<br>Screener (general) | Individual questions asking for the presence of each of following type of delusions:<br>Persecutory/ Jealous<br>Mind reading Thought insertion<br>Thought broadcasting<br>Influence | Individual questions asking for the presence of each of following type of delusions:<br>Persecutory /Jealous<br>Mind reading<br>Thought insertion/ broadcasting/ withdrawal<br>Guilt<br>Grandiose<br>Nihilistic<br>Of Reference<br>Erotomanic<br>Paranormal / Religious | Individual questions asking for the presence of each of following type of delusions:<br>Auditory<br>Tactile |
| **Hallucinations** | Individual questions asking for the presence of each of following type of hallucinations:<br>Any modality(auditory, visual, olfactory, somatic, sexual)<br>Running Commentary<br>Auditory | Individual questions asking for the presence of each of following type of hallucinations:<br>Auditory | Individual questions asking for the presence of each of following type of hallucinations:<br>Visual<br>Auditory<br>Tactile<br>Somatic<br>Gustatory/Olfactory | Individual questions asking for the presence of each of following type of hallucinations:<br>Visual<br>Auditory |

**Supplementary Table 2: Harmonization of Phenotypes Across Cohorts** details the specific wording of the questions used by each cohort's to assess each phenotype. In addition to the 19 selected phenotypes, we include the harmonization of hospitalization (bottom row).

| | GPC | NGAP | PAISA | PUMAS-LATAM |
|---|---|---|---|---|
| **Fatigue** | During that time, did you have as much energy as usual? Did you get exhausted and worn out during the day, even when you weren't working very hard? | Did you feel tired or without energy almost every day? | **SCID:** What has your energy level been like? Tired all the time? Nearly everyday? (Fatigue or loss of energy nearly everyday).<br><br>**Supp. Q:** Have you had a period of at least a few weeks in your life where every night you have felt a decrease in the need to sleep? | During that time, did you have as much energy as usual? Did you get exhausted and worn out during the day, even when you weren't working very hard? |
| **Anhedonia** | Did you ever have a period of time when you were unable to enjoy things as much as usual? For example, taking a walk, spending time with friends, or working at your hobbies or interests? Did this last most of the day, nearly every day for at least one week? | Were you ever much less interested in most things or much less able to enjoy the things you used to enjoy most of the time, for two weeks? | ***SCID:*** During that time, did you lose interest o pleasure in things you usually enjoy? (Diminished interest or pleasure in all, or almost all, activities most of the day, nearly everyday.<br><br>***Supp. Q***: At some point in your life you have had a period of at least a few weeks in which you have felt most of the time a loss of interest in the things you usually enjoy or enjoy doing? | Did you ever have a period of time when you were unable to enjoy things as much as usual? For example, taking a walk, spending time with friends, or working at your hobbies or interests? Did this last most of the day, nearly every day for at least one week? |
| **Suicide Ideation** | | | ***SCID:*** Have you ever wished you were dead? | Did you ever think about killing yourself? Were you thinking about death or dying a lot? Did you think about harming yourself? What happened? |
| **Suicide Attempt** | Have you ever attempted suicide? | | ***SCID:*** Have you ever tried to kill yourself? | Have you ever attempted suicide? |
| **Suicidality** | During that time, did you feel that life was not worth living? Were you thinking about death or dying a lot? Did you think about harming yourself or even made an attempt at suicide? What happened? When was this? How long did you feel like that? Thinking of suicide, wishing to be dead, attempts to kill self, whether depressed or not.<br><br>Merged with Suicide Attempt question | Did you repeatedly think about death, or have any thoughts of killing yourself, or have any intent or plan to kill yourself? Did you attempt suicide? | ***SCID:*** Have things been so bad that you thought a lot about death or that you would be better off dead? Have you thought about taking your own life? Recurrent thoughts of death (not just fear of dying), recurrent suicidal ideation without a specific plan, or a suicide attempt, or a specific plan for committing suicide.<br><br>Suicide Attempt & Ideation combined | During that time, did you feel that life was not worth living? Were you thinking about death or dying a lot? Did you think about harming yourself or even made an attempt at suicide? What happened? When was this? How long did you feel like that? Thinking of suicide, wishing to be dead, attempts to kill self, whether depressed or not.<br><br>Suicide Attempt & Ideation combined |
| **Blunted Affect** | Restricted Affect: Subject's emotional responses are restricted in range and at interview there is an impression of bland indifference. A relatively expressionless face or unchanging facial expression.<br><br>Blunted Affect: A global diminution of emotional response. Subject's emotional responses are persistently flat and show a complete failure to 'resonate' to external change. The difference between restricted and blunted affect should be regarded as one of degree, with 'blunted' only being rated in extreme cases. | | **SCID**: Diminished Emotional Expressiveness--Includes a reduction in the facial expression of emotions, eye contact, speech intonation (prosody) and hand movements, the head and the face that usually gives an emotional emphasis to speech. | Restricted Affect: Subject's emotional responses are restricted in range and at interview there is an impression of bland indifference or "lack of contact". A relatively expressionless face or unchanging facial expression.<br><br>Blunted Affect: A global diminution of emotional response. Subject's emotional responses are persistently flat and show a complete failure to 'resonate' to external change. The difference between restricted and blunted affect should be regarded as one of degree, with 'blunted' only being rated in extreme cases. |
| **Alogia** | Negative formal thought disorder: Blocking: Sudden interruption in speech without reason and then begins again on same or different topic. Poverty of content of speech: Talks freely but so vaguely that little information is given in spite of the number of words used. Restricted quantity of speech: Frequently fails to answer, questions have to be repeated, restricted to minimum necessary. | | | Negative formal thought disorder: Blocking: Sudden interruption in speech without reason and then begins again on same or different topic. Poverty of content of speech: Talks freely but so vaguely that little information is given in spite of the number of words used. Restricted quantity of speech: Frequently fails to answer, questions have to be repeated, restricted to minimum necessary. |
| **Negative Psychotic Symptoms** | Blunted Affect and Alogia combined | Did the participant ever in the past have negative symptoms, e.g. Significant reduction of emotional expression or affective flattening, poverty of speech (alogia) or an inability to initiate or persist in goal-directed activities (avolition)? | Blunted Affect and Avolition combined.<br><br>**SCID**: Has there been a period of time lasting at least several months during which you were not working, studying, or doing much? And a period during which you were unable to perform everyday activities like brushing your teeth or bathing? Has anyone ever said that you were not engaging in these or other daily activities? | Blunted Affect, Alogia, and Avolition combined |

**Supplementary Table 2:** (continued)

| | GPC | NGAP | PAISA | PUMAS-LATAM |
|---|---|---|---|---|
| **Decreased Need of Sleep** | How many days were you sleeping less than usual? Subject sleeps much less but there is no complaint of insomnia. | | **SCID:** Did you sleep less that usual? Were you still feeling rested? **(**Decreased need for sleep (feeling rested despite sleeping less than usual; to be contrasted with insomnia)).<br><br>**Supp. Q:** Have you had a period of at least a few weeks in your life where every night you have felt a decrease in the need to sleep | Have you ever/During that time, did you need far less sleep than usual without feeling tired? |
| **Hypersomnia** | How long did you have problems sleeping more than usual? Subject complains of sleeping at least two hours longer than usual, more or less daily. | | **SCID:** How have you been sleeping? (Trouble falling asleep, waking up frequently, OR sleeping too much? Has it been nearly every night? How many [check if Hypersomnia]<br><br>**Supp. Q:** Have you ever had a period of at least a few weeks in your life where you felt a greater need to sleep every night? (At least during the usual bad hours). | During that time, did you sleep more than usual? How long did you have problems sleeping more than usual? (at least 2 hours) |
| **Sleep Disturbances** | Insomnia and Hypersomnia questions combined | Did you have trouble sleeping nearly every night (difficulty falling asleep, waking up in the middle of the night, early morning waking or sleeping excessively)? | Insomnia and Hypersomnia questions combined | Insomnia and Hypersomnia questions combined |
| **Irritability** | Have you ever found you were easily irritated, that any little problem provoked you, or that other people said you were much too impatient? Did this last most of the day, nearly every day for at least two weeks?<br><br>Irritable mood; Have you ever felt very irritable or excessively annoyed with others, such that you lost your temper often, shouted at people, or even got into fights? Can you describe that feeling? Was it out of character for you? | Have you ever been persistently irritable, for several days, so that you had arguments or verbal or physical fights, or shouted at people outside your family? Have you or others noticed that you have been more irritable or over reacted, compared to other people, even in situations that you felt were justified? | **SCID:** Were you often irritable?<br><br>**SCID:** A distinct period (lasting several days) of abnormally and persistently elevated, expansive, or irritable mood and abnormally and persistently increased activity or energy? (Check if irritable) | Have you ever found you were easily irritated, that any little problem provoked you, or that other people said you were much too impatient? Did this last most of the day, nearly every day for at least two weeks?<br><br>Irritable mood; Have you ever felt very irritable or excessively annoyed with others, such that you lost your temper often, shouted at people, or even got into fights? Can you describe that feeling? Was it out of character for you? |
| **Flight of Ideas** | Racing thoughts: During that time, did you find your thoughts crowding into or racing through your mind, so that you felt like you couldn't keep up with them? Can you describe what that felt like? How long were you feeling like that? Subject experiences thoughts racing through his/her head or others observe flight of ideas that make it difficult to follow what subject is talking about. | Notice your thoughts going very fast or running together or racing or moving very quickly from one subject to another? | **SCID:** Did you have thoughts racing through your head? Flight of ideas or subjective experience that thoughts are racing<br><br>**Supp. Q:** Have you had a period of at least 1 week at any point in your life when you felt like your mind was racing, full of more ideas and thoughts, compared to the usual? | Racing thoughts: During that time, did you find your thoughts crowding into or racing through your mind, so that you felt like you couldn't keep up with them? Can you describe what that felt like? How long were you feeling like that? Subject experiences thoughts racing through his/her head or others observe flight of ideas that make it difficult to follow what subject is talking about |
| **Hospitalizations** | | Were you hospitalized for these problems? | Have you ever been a patient in a psychiatric hospital? How many times? | How many times have the patient been hospitalized for psychiatric illness? |

**Supplementary Table 2:** (continued)

| Phenotype | Diagnosis Group | Frequency (count) | Total | Proportion |
|---|---|---|---|---|
| Lifetime Depressive Episode | BP-I | 11081 | 14969 | 0.74 |
| | Other BP | 1711 | 2463 | 0.695 |
| | SZA | 2550 | 3268 | 0.78 |
| | SCZ | 9456 | 21094 | 0.448 |
| | MDD | 2805 | 2816 | 0.996 |
| Anhedonia | BP-I | 11319 | 15169 | 0.746 |
| | Other BP | 2059 | 2743 | 0.751 |
| | SZA | 2586 | 3315 | 0.78 |
| | SCZ | 10608 | 21398 | 0.496 |
| | MDD | 2761 | 2816 | 0.98 |
| Suicide Ideation | BP-I | 3472 | 6280 | 0.553 |
| | Other BP | 1098 | 1431 | 0.767 |
| | SZA | 382 | 707 | 0.54 |
| | SCZ | 1490 | 4548 | 0.328 |
| | MDD | 2687 | 2816 | 0.954 |
| Suicide Attempt | BP-I | 2303 | 6447 | 0.357 |
| | Other BP | 626 | 1445 | 0.433 |
| | SZA | 336 | 831 | 0.404 |
| | SCZ | 1100 | 4661 | 0.236 |
| | MDD | 1410 | 2816 | 0.501 |
| Suicidality | BP-I | 8156 | 15193 | 0.537 |
| | Other BP | 1595 | 2747 | 0.581 |
| | SZA | 2092 | 3334 | 0.627 |
| | SCZ | 7358 | 21660 | 0.34 |
| | MDD | 2688 | 2816 | 0.955 |
| Fatigue | BP-I | 10536 | 15038 | 0.701 |
| | Other BP | 1962 | 2720 | 0.721 |
| | SZA | 2509 | 3241 | 0.774 |
| | SCZ | 9399 | 19410 | 0.484 |
| | MDD | 2640 | 2816 | 0.938 |
| Hypersomnia | BP-I | 3355 | 8580 | 0.391 |
| | Other BP | 635 | 1666 | 0.381 |
| | SZA | 944 | 2514 | 0.375 |
| | SCZ | 2337 | 9869 | 0.237 |
| | MDD | 829 | 2816 | 0.294 |
| Sleep Disturbances | BP-I | 11462 | 15160 | 0.756 |
| | Other BP | 2042 | 2747 | 0.743 |
| | SZA | 2533 | 3331 | 0.76 |
| | SCZ | 10434 | 21646 | 0.482 |
| | MDD | 2745 | 2816 | 0.975 |
| Irritability | BP-I | 13491 | 15193 | 0.888 |
| | Other BP | 1534 | 2661 | 0.576 |
| | SZA | 2677 | 3281 | 0.816 |
| | SCZ | 9527 | 20987 | 0.454 |
| | MDD | 423 | 2436 | 0.174 |

| Phenotype | Diagnosis Group | Frequency (count) | Total | Proportion |
|---|---|---|---|---|
| Decreased Need of Sleep | BP-I | 13798 | 14644 | 0.942 |
| | Other BP | 1514 | 2599 | 0.583 |
| | SZA | 2234 | 3235 | 0.691 |
| | SCZ | 6835 | 20432 | 0.335 |
| | MDD | 340 | 2762 | 0.123 |
| Flight Ideas | BP-I | 14089 | 15187 | 0.928 |
| | Other BP | 1603 | 2739 | 0.585 |
| | SZA | 2413 | 3329 | 0.725 |
| | SCZ | 6447 | 21591 | 0.299 |
| | MDD | 259 | 2762 | 0.094 |
| Lifetime Manic Episode | BP-I | 15193 | 15193 | 1 |
| | Other BP | 0 | 0 | 0 |
| | SZA | 2241 | 3037 | 0.738 |
| | SCZ | 6360 | 21200 | 0.3 |
| | MDD | 2 | 2816 | 0.001 |
| Grandiosity | BP-I | 11407 | 15193 | 0.751 |
| | Other BP | 1133 | 2747 | 0.412 |
| | SZA | 1869 | 3333 | 0.561 |
| | SCZ | 6532 | 21660 | 0.302 |
| | MDD | 81 | 2816 | 0.029 |
| Delusions | BP-I | 11902 | 15184 | 0.784 |
| | Other BP | 1423 | 2745 | 0.518 |
| | SZA | 3135 | 3334 | 0.94 |
| | SCZ | 20280 | 21657 | 0.936 |
| | MDD | 266 | 2811 | 0.095 |
| Hallucinations | BP-I | 9698 | 15193 | 0.638 |
| | Other BP | 1244 | 2747 | 0.453 |
| | SZA | 3044 | 3334 | 0.913 |
| | SCZ | 19433 | 21661 | 0.897 |
| | MDD | 474 | 2816 | 0.168 |
| Positive Psychotic Symptoms | BP-I | 12869 | 15193 | 0.847 |
| | Other BP | 1682 | 2747 | 0.612 |
| | SZA | 3275 | 3334 | 0.982 |
| | SCZ | 21153 | 21661 | 0.977 |
| | MDD | 559 | 2816 | 0.199 |
| Blunted Affect | BP-I | 1176 | 8662 | 0.136 |
| | Other BP | 146 | 1679 | 0.087 |
| | SZA | 1057 | 2548 | 0.415 |
| | SCZ | 6437 | 10251 | 0.628 |
| | MDD | 17 | 2814 | 0.006 |
| Alogia | BP-I | 378 | 6272 | 0.06 |
| | Other BP | 74 | 743 | 0.1 |
| | SZA | 579 | 2226 | 0.26 |
| | SCZ | 4394 | 9051 | 0.485 |
| | MDD | NA | NA | NA |
| Negative Psychotic Symptoms | BP-I | 3414 | 15193 | 0.225 |
| | Other BP | 479 | 2747 | 0.174 |
| | SZA | 1709 | 3333 | 0.513 |
| | SCZ | 14090 | 21659 | 0.651 |
| | MDD | 36 | 2815 | 0.013 |

**Supplementary Table 3. Phenotype Endorsement proportion by Diagnosis Group** (BP-I, Other BP, SZA, SCZ, and MDD).

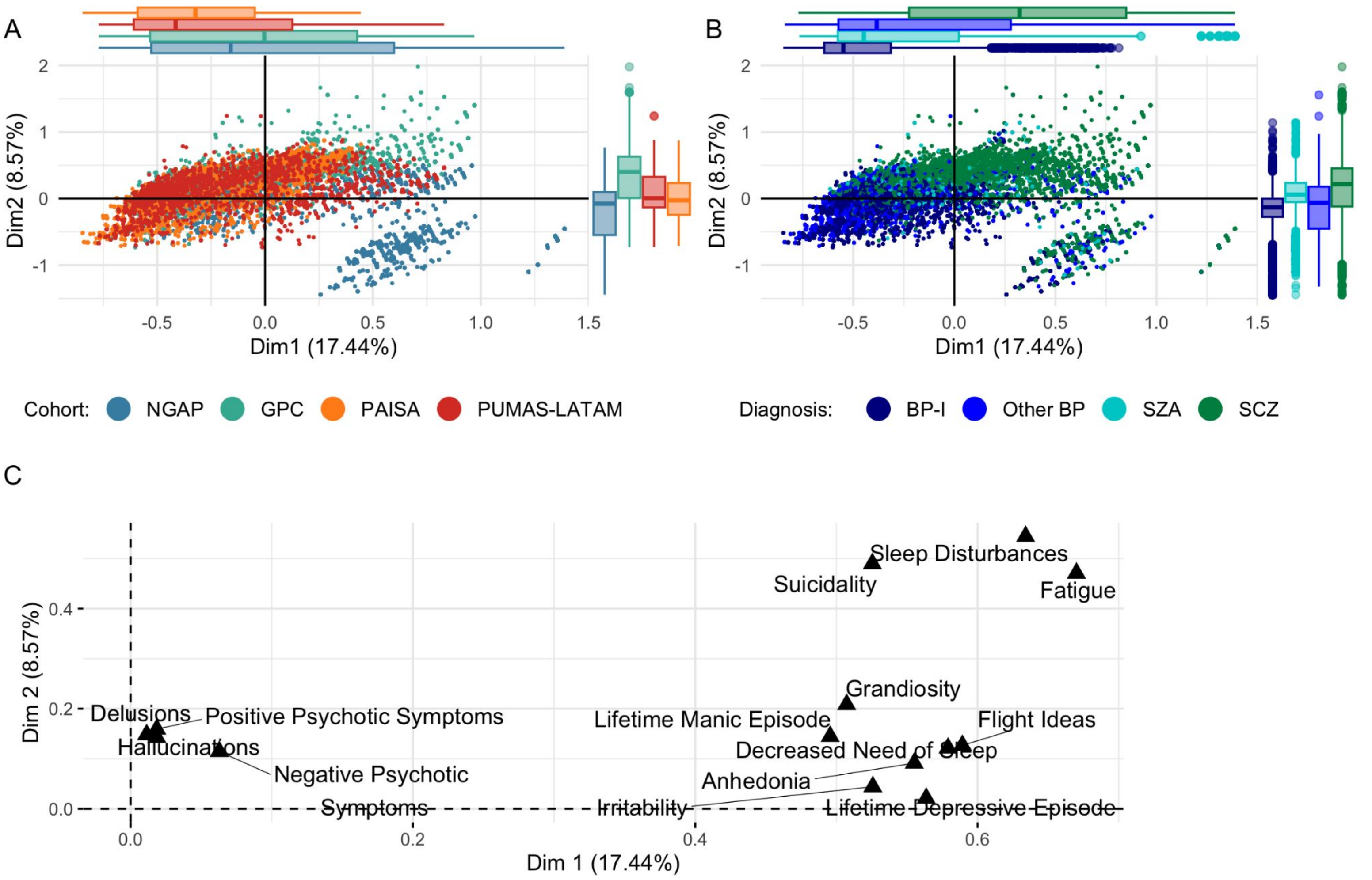


**Supplementary Figure 1. PUMAS-wide Multiple Correspondence Analysis (MCA) excluding MDD patients (A & B)** depict the first two dimensions (Dim1 and Dim2) of an MCA performed using item-level phenotypes assessed in all cohorts (positive psychotic symptoms, delusions, hallucinations, negative psychotic symptoms, irritability, flight ideas, grandiosity, lifetime manic episode, lifetime depressive episode, suicidality, anhedonia, sleep disturbances, decreased need of sleep, and fatigue). Individuals are colored by cohort (NGAP, GPC, PAISA, PUMAS-LATAM (**A**) and by Diagnosis (BP-I, Other BP, SZA, and SCZ) (**B**). The percentage of variation (inertia) explained by each dimension is indicated in parentheses. The marginal box plots summarize the distribution of the MCA scores along Dim1 and Dim2 for each cohort or diagnosis group. The central line in each box represents the median of the data, while the hinges of the box indicate the first and third quartiles. (**C**) Visualizes the variable correlations to the MCA dimensions. MDD patients are excluded

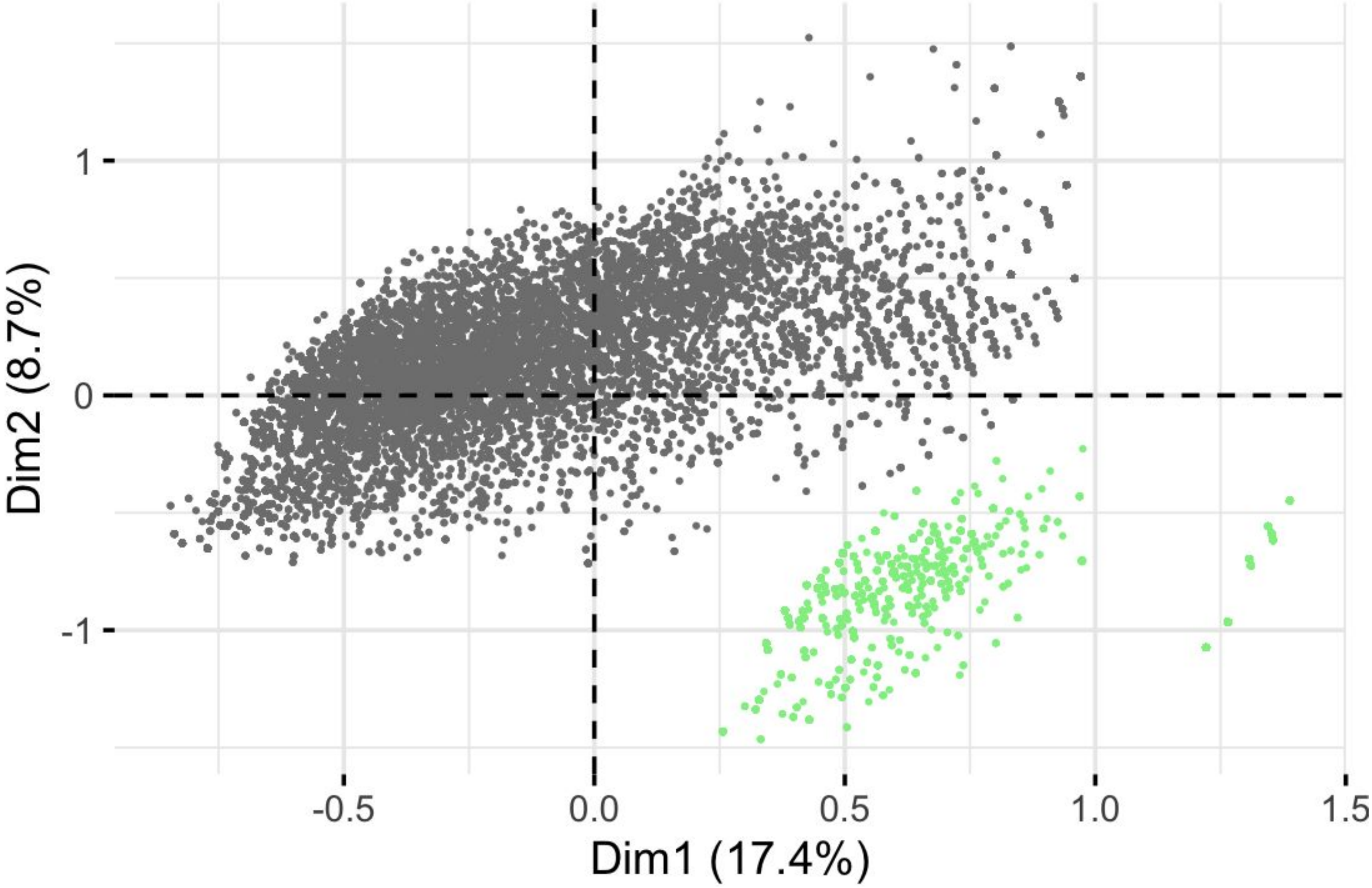


**Supplementary Figure 2. PUMAS-wide Multiple Correspondence Analysis (MCA) excluding MDD patients** highlighting in green NGAP participants cluster that branched out of MINI depression module .

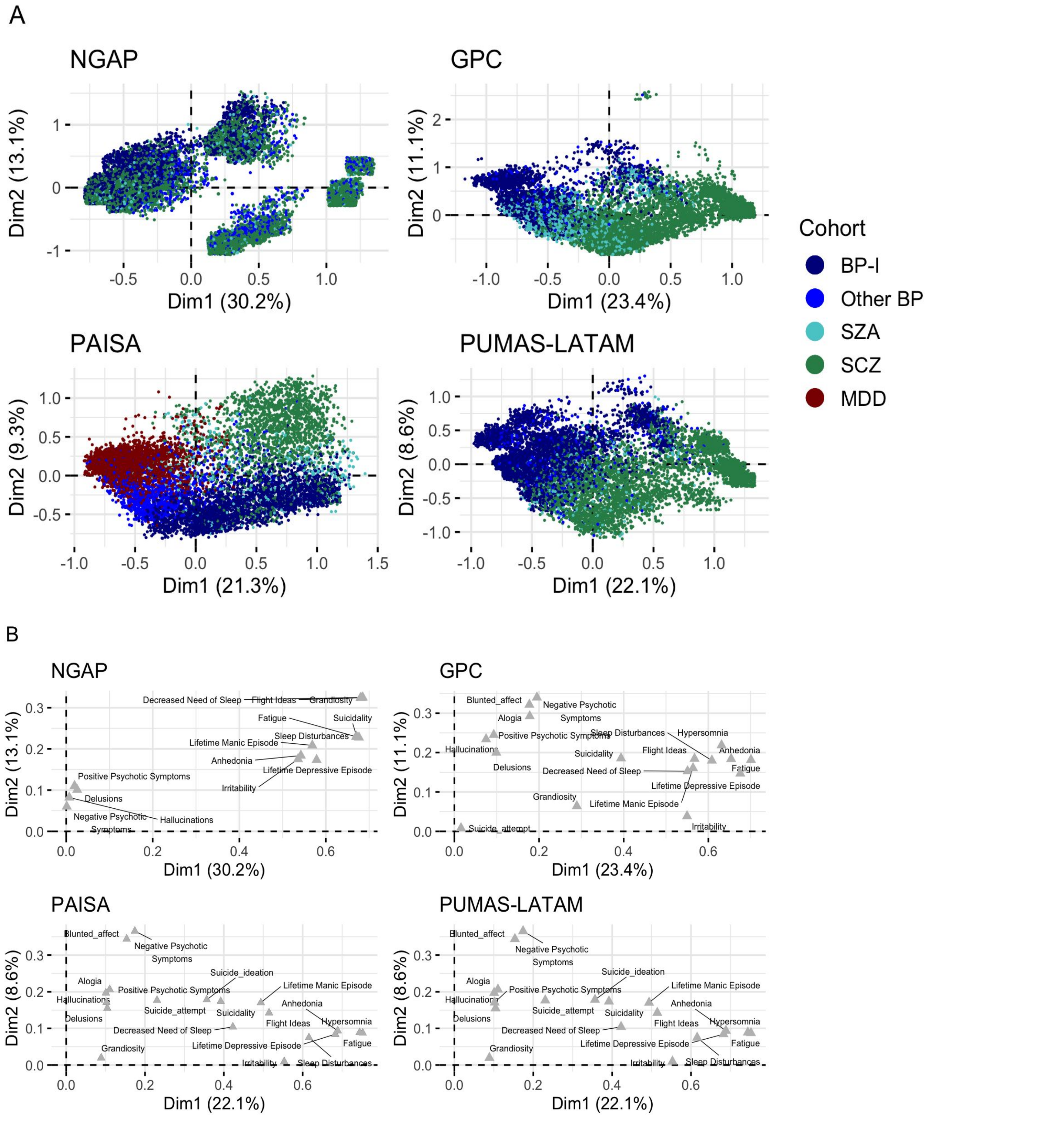


**Supplementary Figure 3. Cohort-specific Multiple Correspondence Analysis (MCA)** (**A**) displays the distribution of individuals across the first two dimensions (Dim1 and Dim2) of an MCA performed using item-level phenotypes available for each Cohort colored by diagnosis (BP-I, Other BP, SZA, SCZ, MDD). The percentage of variation (inertia) explained by each dimension is indicated in parentheses. (**B**) Illustrates the variable correlations to the dimensions for the MCA.

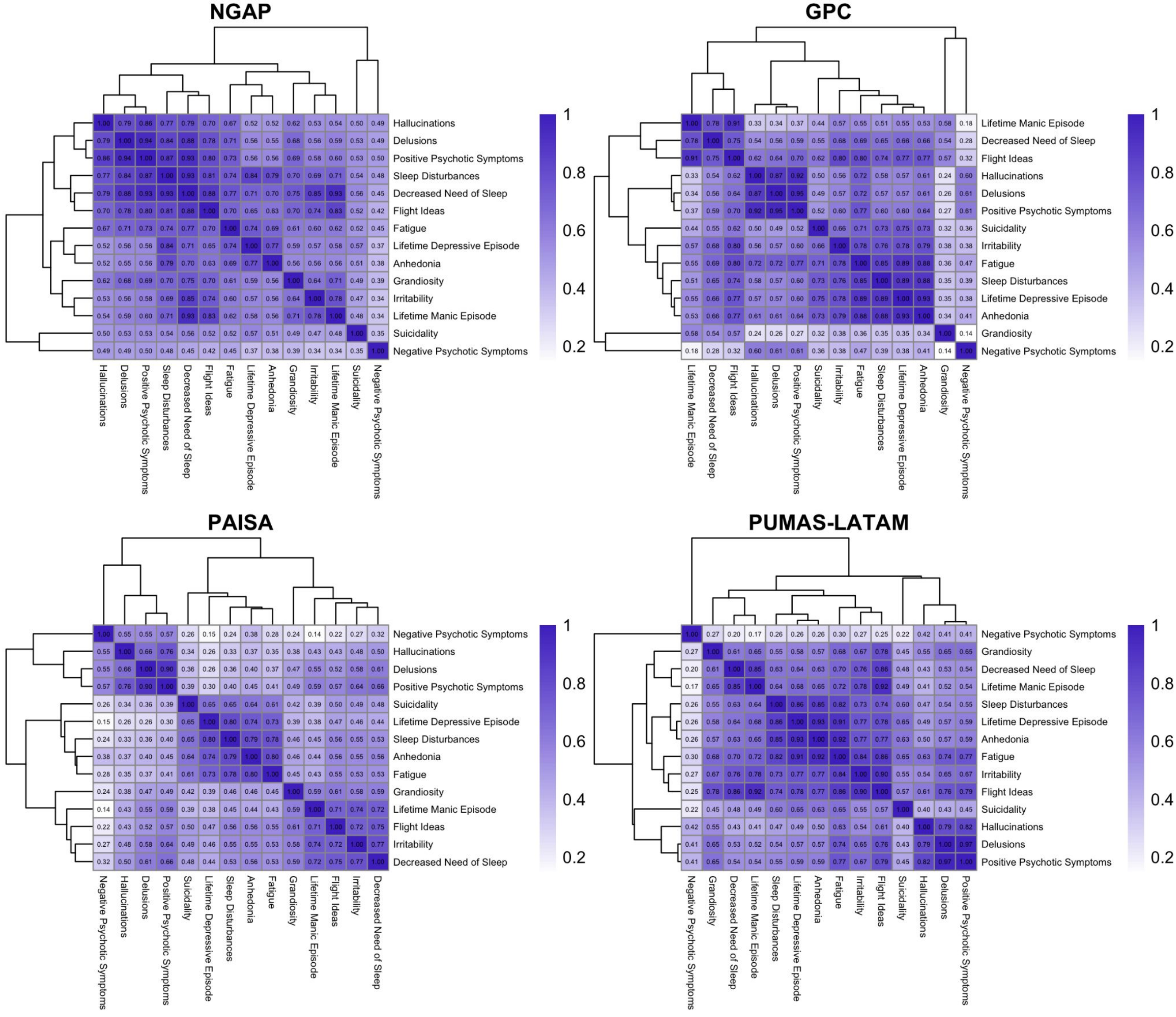


**Supplementary Figure 4. Jaccard Similarity Matrix of Phenotypic Variables for each PUMAS Cohort (excluding MDD patients from Paisa Cohort)** Each cell in the matrix corresponds to the Jaccard's Similarity of two phenotypes, calculated as the proportion of instances where both phenotypes are endorsed relative to the total number of instances where at least one of the phenotypes is endorsed. The gradient scale from light to dark reflects the range of Jaccard Similarity Scores, from 0 (no similarity) to 1 (perfect similarity).

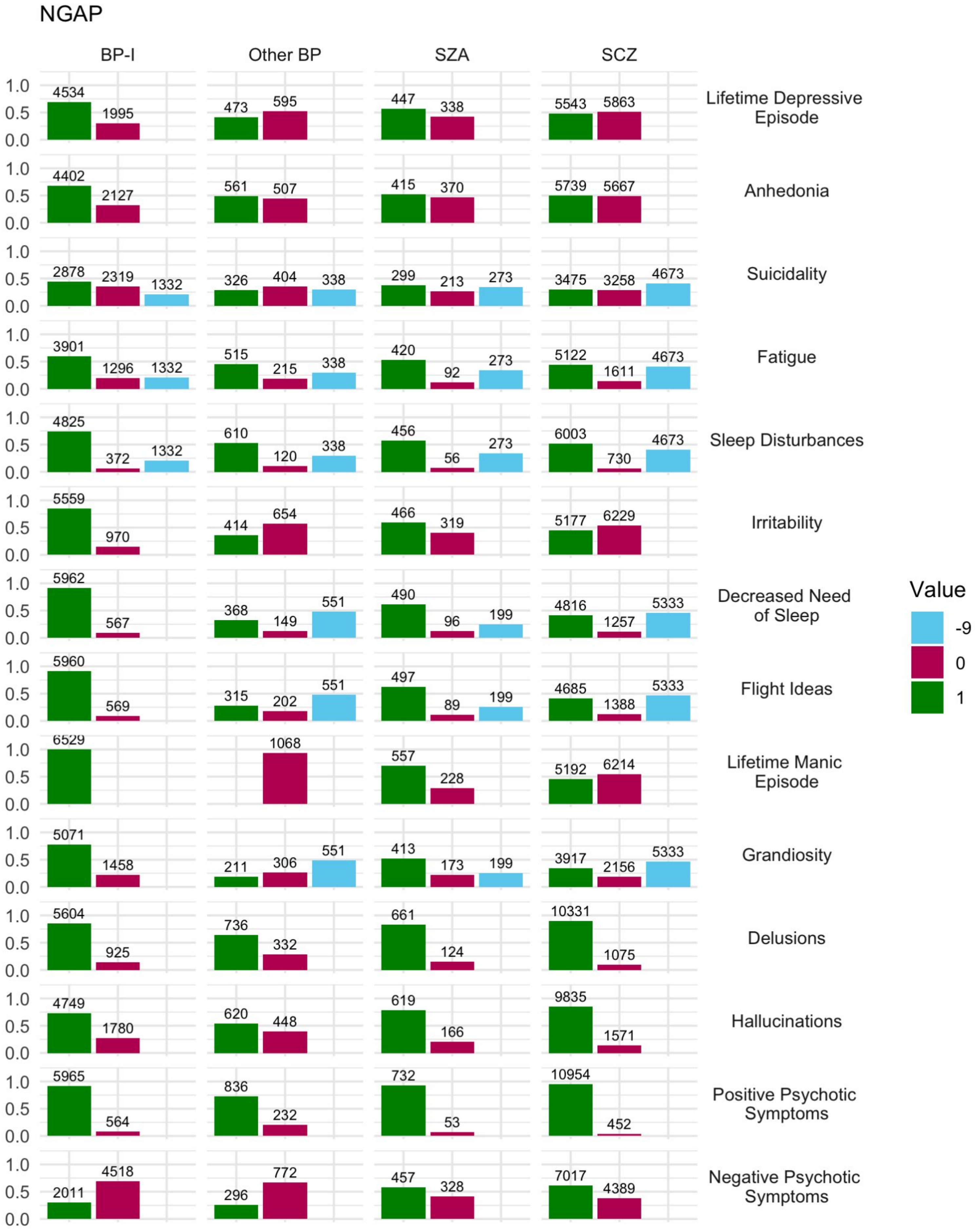


**Supplementary Figure 5A Phenotype Endorsement including branching for NGAP cases** displaying branching logic across diagnostic groups (BP-I, Other BP, SZA, and SCZ) where 1= endorsed, 0= not endorsed, -9=skipped due to branching logic of instrument, and NA = missing values.

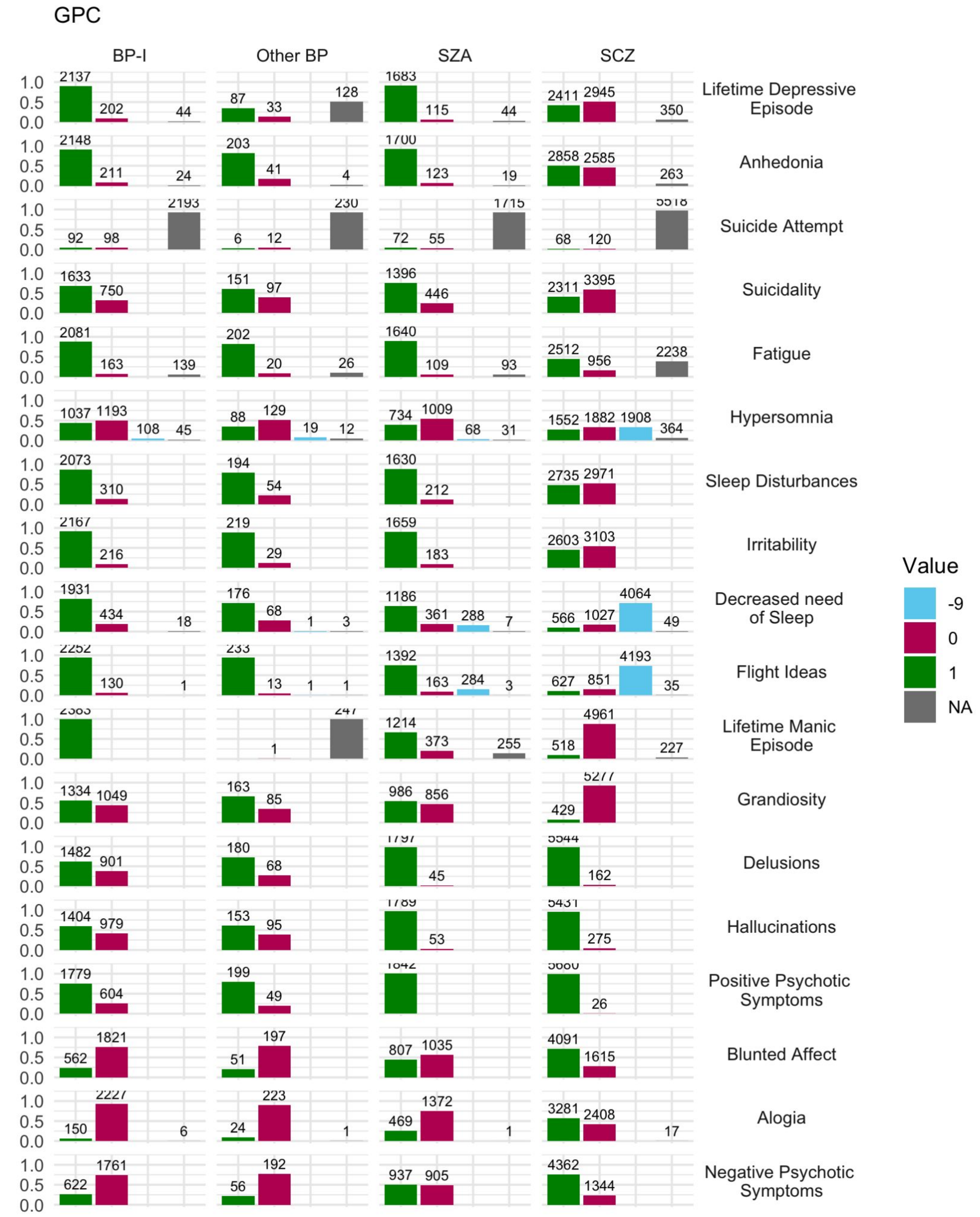


**Supplementary Figure 5B Phenotype Endorsement including branching for GPC cases** displaying branching logic across diagnostic groups (BP-I, Other BP, SZA, and SCZ) where 1= endorsed, 0= not endorsed, -9=skipped due to branching logic of instrument, and NA = missing values.

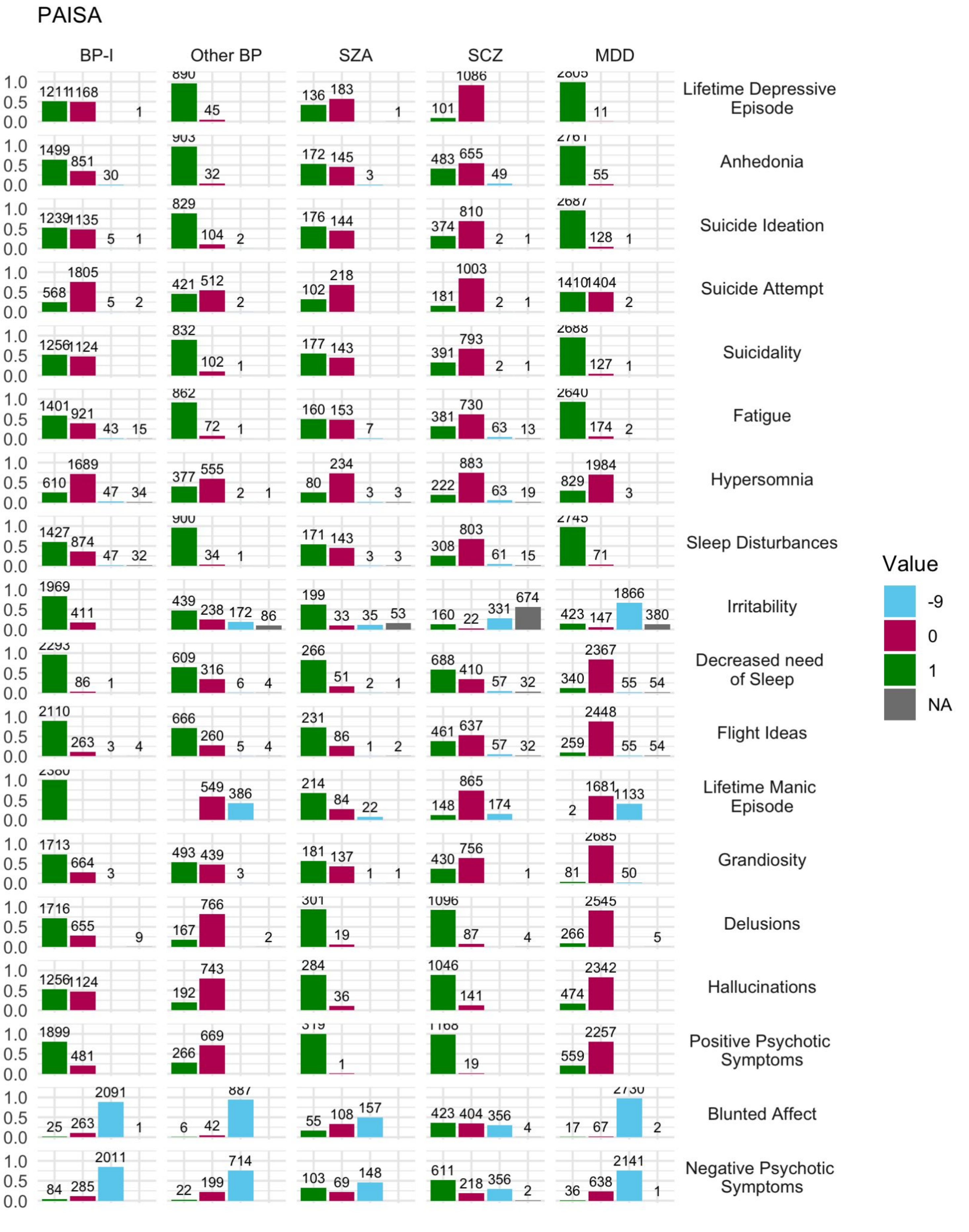


**Supplementary Figure 5C Phenotype Endorsement including branching for PAISA cases** displaying branching logic across diagnostic groups (BP-I, Other BP, SZA, SCZ, and MDD) where 1= endorsed, 0= not endorsed, -9=skipped due to branching logic of instrument, and NA = missing values.

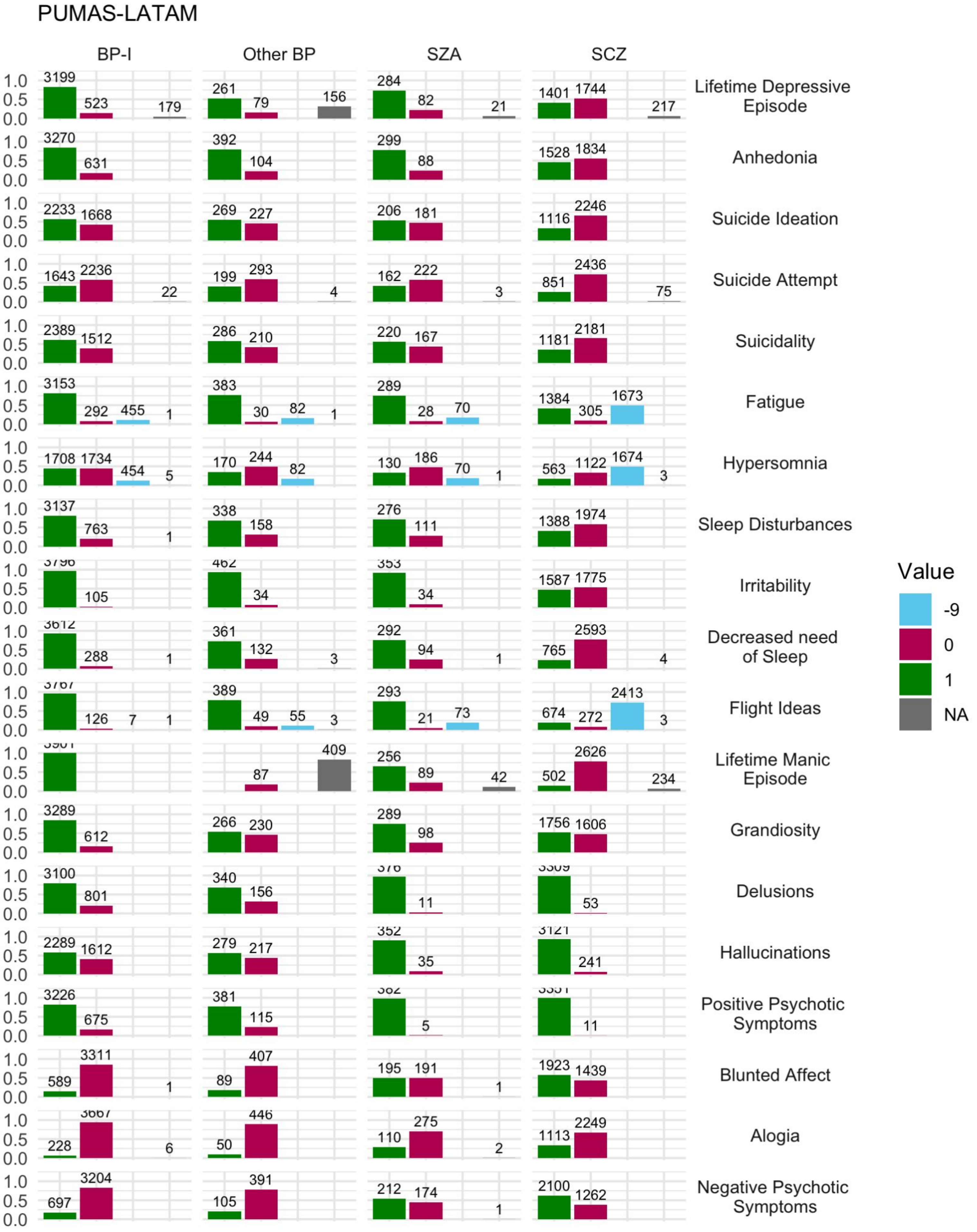


**Supplementary Figure 5D Phenotype Endorsement including branching for PUMAS-LATAM cases** displaying branching logic across diagnostic groups (BP-I, Other BP, SZA, and SCZ) where 1= endorsed, 0= not endorsed, -9=skipped due to branching logic of instrument, and NA = missing values.
